# Volatility-Level Inference Indexes Psychosis Spectrum Symptoms Independent of Age in Transdiagnostic Help-Seeking Youth

**DOI:** 10.64898/2026.06.24.26356447

**Authors:** Milad Soltanzadeh, Stephanie H. Ameis, Colleen E. Charlton, Kristin Cleverley, Darren B. Courtney, Erin W. Dickie, Daniel Felsky, George Foussias, Benjamin Goldstein, John D. Griffiths, Nicole Kozloff, Denisa Lazar, Aloha Narajos, Yuliya Nikolova, Oreoluwa A. Ogundipe, Thalia Phi, Alexia Polillo, Connie Putterman, Lena C. Quilty, Diya Shah, Aristotle N. Voineskos, Wei Wang, Zheng Wang, Andreea O. Diaconescu, TAY CAMH Cohort Study Group

**Affiliations:** Institute of Medical Science, University of Toronto, Toronto, Ontario, Canada; Krembil Centre for Neuroinformatics, Centre for Addiction and Mental Health, Toronto, Ontario, Canada; Campbell Family Mental Health Research Institute, Centre for Addiction and Mental Health, Toronto, Ontario, Canada; Cundill Centre for Child and Youth Depression, Campbell Family Mental Health Research Institute, Centre for Addiction and Mental Health, Toronto, Ontario, Canada; Temerty Faculty of Medicine, Department of Psychiatry, University of Toronto, Toronto, Ontario; Margaret and Wallace McCain Centre for Child, Youth and Family Mental Health, Centre for Addiction and Mental Health, Toronto, Ontario, Canada; Centre for Addiction and Mental Health, Toronto, Ontario, Canada

**Author notes:** **Corresponding author: Email:**, **Address:** 12th floor, Centre for Addiction and Mental Health, 250 College St., Toronto, ON, Canada M5T 1R8. all authors except for the first and the last author contributed equally and they are listed in alphabetical order.

**Keywords:** mismatch negativity, psychosis spectrum symptoms, precision-weighted prediction errors, hierarchical Gaussian filter, predictive coding, EEG, computational psychiatry, adolescents

## Abstract

**Background:** Psychosis spectrum symptoms (PSS) are prevalent in youth and are associated with increased risk for psychotic disorder, suicidality, and functional impairment. Computationally, PSS may stem from altered predictive coding of basic sensory surprises and environmental volatility. Formalized as hierarchical precision-weighted prediction errors (pwPEs), this altered processing is a proposed mechanistic substrate of aberrant perceptual inference across disorders, including psychosis-risk populations. While the auditory mismatch negativity (MMN) provides an electrophysiological index of pwPEs, it remains unknown if distinct hierarchical pwPE components distinguish youth who endorse PSS.

**Methods:** A sample of 131 participants (PSS–=66, PSS+=65; ages 11–24) from the ongoing Toronto Adolescent and Youth (TAY-CAMH) Cohort study were stratified by PSS status using the PRIME Screen–Revised and were assessed for their psychosocial functioning. 64-channel EEG was recorded during an auditory oddball paradigm with stable and volatile phases. A hierarchical Bayesian model applied to the stimulus stream generated trajectories of low-level sensory and high-level volatility-related pwPEs. Alongside standard phase-averaged event-related potentials (ERPs), Bayesian trajectories derived model-based ERPs.

**Results:** Replicating prior findings in non-clinical controls, stable-phase MMN significantly exceeds volatile-phase MMN and lower psychosocial functioning was associated with reduced volatile-phase MMN amplitude. Age significantly modulated oddball MMN and unweighted prediction errors (δ_1_, δ_2_). Group differences between PSS+ and PSS– were statistically significant for volatility-level pwPE (ε_3_), peaking at ∼180 ms Peri-Stimulus Time (p_FWE-peak_ =.024).

**Conclusions:** Independent of age-related developmental effects, volatility-level pwPE learning (ε_3_) constitutes a more sensitive EEG marker associated withPSS status in help-seeking youth than low-level sensory pwPE.

## Introduction

### Psychosis Spectrum Symptoms in Help-Seeking Youth

Psychosis spectrum symptoms (PSS)—encompassing attenuated perceptual disturbances, reality distortions, and suspiciousness—carry substantial burdens in youth and adolescents (1,2). Critically, PSS are not rare or diagnostically specific: Population prevalence estimates indicate that approximately 17% of children and 7.5% of adolescents endorse psychosis-spectrum experiences, with 10% of young adults reporting delusion-like experiences or hallucinations (3,4). While psychotic symptoms are characteristic of schizophrenia spectrum disorders and clinical high risk for psychosis (CHR), PSS also occur across a wide range of psychiatric and neurodevelopmental conditions and are best understood as a transdiagnostic marker of mental health burden rather than a prodrome of any single disorder (2,4,5).

The clinical burden of PSS in help-seeking youth extends well beyond psychosis risk per se (6,7). Among their documented consequences, PSS confer elevated risk for a range of adverse outcomes—including suicidal ideation, functional impairment, and, in a subset of individuals, progression toward psychosis (1,2,8). Preliminary baseline data from mental health help seeking youth characterized by the Toronto Adolescent and Youth Cohort at the Centre for Addiction and Mental Health (TAY-CAMH) demonstrate that PSS+ youth are substantially more impaired than PSS– peers across global functioning, show elevated suicidality and have extensive diagnostic multimorbidity. Crucially, PSS+ status in the TAY-CAMH Cohort appears to delineate a transdiagnostic threshold of psychiatric severity and not necessarily a psychosis-specific risk group (9). This distinction has direct implications for the interpretation of neural computational markers examined in the present study.

PSS are dimensionally heterogeneous. Applying an exploratory bifactor model to the PSS scores allows decomposition of total variance into a general PSS factor and orthogonal specific factors capturing distinct symptom clusters (10,11). Whether distinct EEG signatures relate differentially to the general versus specific PSS dimensions is unknown.

### The Auditory Mismatch Negativity as a Psychosis Biomarker

As an event-related potential (ERP), the MMN is a difference wave elicited by unpredictable (‘deviant’) stimuli relative to predictable (‘standard’) stimuli (12). By manipulating tone probabilities into stable (constant) and volatile (frequently alternating) phases, the modified oddball paradigm captures how the brain continuously models sensory statistics and environmental volatility (13–15). Two principal mechanistic accounts of MMN generation have been proposed: (i) the adaptation hypothesis, emphasizing habituated neural responses to repeated stimuli (16,17); and (ii) the model-adjustment hypothesis, emphasizing hierarchical refinement of auditory predictions (18).

The most robust change in the MMN amplitude in the literature is a significant reduction observed in chronic schizophrenia (19–23). MMN amplitude reduction is also observed in individuals at CHR for psychosis, with reductions scaling with proximity to psychotic disorder onset (24). Similar MMN alterations are reported across other clinical conditions, including major depressive disorder (MDD), neurodevelopmental disorders (e.g., ADHD and autism), and post-traumatic stress disorder (PTSD) (25–29). While most of these conditions are characterized by reduced MMN amplitude, increases have been observed in PTSD, as well as in adults with autism spectrum disorder relative to children (27). Critically, the direction and magnitude of these alterations also depend on the type of auditory deviant (28), underscoring how inconsistent raw MMN amplitude is as a transdiagnostic marker.

Predictive coding specifies the process MMN indexes (i.e., reconciling the adaptation and model-adjustment accounts (18,30,31)) as a precision-weighted prediction error (pwPE). Computationally, a prediction error signals that sensory input has violated expectation and drives belief updating, weighted by the precision (reliability) of the input relative to prior beliefs. Neurophysiologically, these errors are conveyed by bottom-up glutamatergic signalling along forward connections, whereas predictions descend via backward connections mediated by AMPA and NMDA receptors, respectively (32–35). Precision is encoded as the gain of the neuronal populations broadcasting the error — a modulatory, NMDAR–dependent mechanism (36). The MMN thus offers a non-invasive index of this hierarchical inferential signalling.

### Computational Framework: Hierarchical Bayesian Learning and pwPE

Building on the predictive coding theory, our computational modelling approach employs the Hierarchical Gaussian Filter (HGF) (37,38), a framework in which an observer’s beliefs are organized hierarchically and updated via pwPE on each trial. HGF captures how an individual hierarchically updates their beliefs by tracking low-level sensory prediction errors, high-level environmental volatility, and multi-level uncertainty.

Prior work from our group demonstrated that increased low-level sensory pwPE learning is associated with conversion to psychosis in CHR individuals, and is detectable in help-seeking youth (39,40). Under HGF framework, MMN reductions in psychosis-risk populations reflect enhanced ε_2_ learning, consistent with primary deficits in NMDAR glutamate synaptic plasticity in layer-3 pyramidal neurons in prefrontal cortex, leading to cortical disinhibition and increased sensory excitability (41–44). On the other hand, enhanced perception of environmental volatility (indexed by ε_3_) constitutes a distinct, higher-level mechanism. Our prior work demonstrated increased volatility perception during social learning in FEP patients (45). Aberrant volatility signaling has also been proposed to underlie other clinical conditions such as autism spectrum disorder and anxiety (46,47).

Crucially, the pharmacological specificity of ε_3_ has been established by Weber, Diaconescu et al. (48), who demonstrated that NMDAR blockade selectively reduces the EEG expression of ε_3_ — but leaves ε_2_ statistically intact. This pharmacological dissociation directly motivates our prediction that ε_3_, rather than ε_2_, would index PSS-associated alterations in hierarchical auditory inference. A normative baseline for this paradigm in healthy individuals has been established by Charlton et al. (14), who applied the identical stable/volatile oddball paradigm with HGF modelling to 43 non-clinical controls, demonstrating robust ε_3_ expression and a specific association between volatile-phase MMN and social functioning.

### Aims and Hypotheses

The present study addresses the following specific aims in a subset of youth collected as part of the ongoing TAY-CAMH Cohort Study of mental health treatment-seeking youth with a variety of clinical diagnoses and presentations: **Aim 1.** Characterize the overall MMN effect and its modulation by auditory task context (stable vs. volatile probability phases); **Aim 2.** Test whether PSS+ youth exhibit altered ERP-level and model-based EEG signatures (ε_2_, ε_3_) relative to PSS– youth; **Aim 3.** Examine the dimensional specificity of pwPE–derived difference waveform associations by relating model-based ERPs to the general and specific factors from bifactor analysis of the PRIME-R.

We hypothesized that: **H1.** we would observe competing directional effects for MMN amplitude in PSS+ youth: primarily, assuming linear scaling with proximity to psychosis, we anticipated reduced MMN amplitudes; alternatively, we speculated that elevated comorbid anxiety or PTSD might drive an increase in MMN amplitude; **H2.** ε_3_ would distinguish PSS groups more robustly than ε_2_, given its higher-order representational role; **H3.** volatility-level parameters (δ_2_, ψ_3_) would show dimensional associations with the general PSS factor, theoretically reflecting a transdiagnostic mechanism of reduced hierarchical inference; and **H4.** the volatile-phase MMN would specifically associate with GF:Social scores, replicating the phase-specific functioning association previously reported by Charlton et al. (14).

## Materials and Methods

### Study Design and Setting

This study constitutes a nested sub-study within the ongoing TAY-CAMH Cohort Study, a multimodal, hospital-based longitudinal study of up to 1500 children and youth (ages 11–24) presenting to the CAMH for mental health care services and followed over five years. The TAY-CAMH Cohort Study launched in 2021 and has recruited nearly 90% of the total sample to date (9). The current analysis employs a prospective mixed design and uses a subset of the available TAY data from baseline, 6-month assessments (including blood biomarkers, neurocognitive, and clinical measures) and 1-year follow-up neuroimaging measurements (49). The present analyses report on cross-sectional baseline EEG data. This study was approved by the CAMH Research Ethics Board in compliance with the World Medical Association Declaration of Helsinki.

### Participants

Participants were recruited from the TAY-CAMH Cohort and PSS were measured using the PRIME-R, a 12-item self report assessment (see Section S1 for full list) scored on a 0–6 Likert scale (50). PSS+ and PSS– group classification was based on the Extreme Agreement Index (EAI) criterion: endorsement of >1 item at the maximum response (“definitely agree,” score 6) or >3 items at “somewhat agree” (score 5) (50,51). This threshold-based dichotomisation is distinct from the PRIME-R total score, which was used separately as a continuous covariate in dimensional analyses. In the broader TAY-CAMH Cohort, 49.2% of youth completing the PRIME-R met EAI criteria for PSS (9). This level of PSS endorsement is likely consistent with the help-seeking nature of the sample. Participants were generally recruited to complete EEG only if they declined or were ineligible for MRI. After EEG quality control, the final analytic sample available for the current analysis comprised N=131 participants (PSS–: n=66; PSS+: n=65).

In addition to PRIME-R, participants underwent a series of clinical assessments to evaluate their psychiatric disorders and psychosocial functioning. The PRIME-R comprises 12 items organised into five domains inspired by the original validation work (52): Unusual Thought Content/delusional mood (UTC; items 1, 3, 5, 6 and 9), Grandiosity/Superstitions (items 2, 4 and 8), Persecutory Ideation (item 7) and Auditory Hallucinations (items 10–11). Item 12 (“going crazy”) constitutes a standalone metacognitive distress item. These a priori subscale groupings provided the interpretive framework for labelling the specific factors emerging from bifactor analysis.

### Bifactor Analysis of the PRIME-R

An exploratory bifactor analysis (EFA) approach utilizing the Schmid-Leiman orthogonalization (53) transformation was adopted. The model was estimated in Python using the *FactorAnalyzer* (*v0.5.1;* https://factor-analyzer.readthedocs.io/en/latest/index.html) module. The optimal number of specific underlying factors was determined empirically via visual inspection of the scree plot (retention of eigenvalues > 1) (54). The Schmid-Leiman transformation was then applied and extracted one general PSS factor and two orthogonal specific factors (Specific_1 and Specific_2). Factor scores were extracted for subsequent covariate analyses.

### Auditory Oddball Paradigm

The auditory MMN paradigm consisted of a train of binaural pure sinusoidal tones (528 Hz vs. 440 Hz). Participants passively listened via wired in-ear headphones while performing a visual distraction (55). Following the design by Weber et al. (15), tone probabilities systematically fluctuated between stable phases (constant probabilities) and volatile phases (frequently alternating probabilities; see Figure 1.A and Table 1 for more details). To present the auditory and visual stimuli, we employed *PsychToolbox* (*PTB3, version 3.0.14*; psychotoolbox.org) (56) running within *MATLAB (R2018a)*.

**Figure 1.**
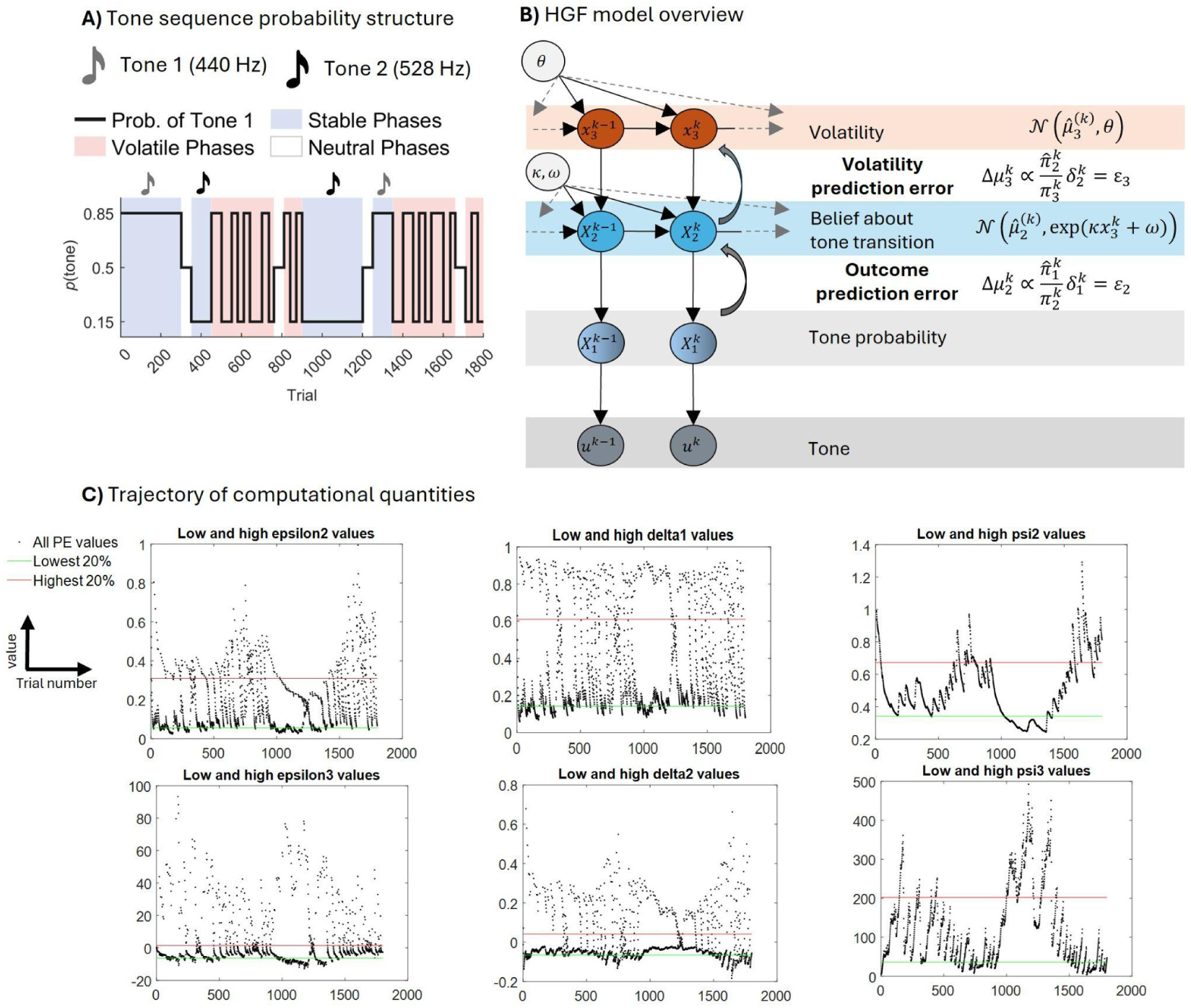
Probability structure of the oddball MMN paradigm and HGF model overview. **A)** The oddball MMN paradigm consists of a tone sequence (440 Hz and 528 Hz). The sequence fluctuates between stable phases (constant probability for ≥ 100 trials) and volatile phases (transitions every 25–60 trials). The plotted line represents the probability of Tone 1 (*p*_1_). Standard and deviant roles are assigned based on the relative probability of the two tones (*p*_1_ vs. 1-*p_1_*) within each stable period. **B)** A three-level Hierarchical Gaussian Filter (HGF) is employed to model the subject’s belief updating process. Level 1 (*X ^k^*): Represents the categorical outcome of trial *k*, level 2 (*X ^k^*): Tracks the evolving belief regarding tone transition probabilities at trial *k* and level 3 (*x ^k^*): Captures the environmental volatility at trial *k*, or the rate at which the transition probabilities at Level 2 are expected to change. The model updates through a reciprocal process where bottom-up prediction errors (outcome and volatility PEs) drive state updates, while top-down parameters (ɵ, κ and ω) govern the coupling between levels and the individual’s learning rate. **C)** Trajectory of precisions and prediction errors for the Bayes optimal agent simulated to reflect the temporal fluctuations during the pre-defined oddball task.

**Table 1.** Experimental paradigm, Preprocessing and QC Specifications. Hz: Hertz; ICA: Independent Component Analysis; mm: Millimeters; ms: Milliseconds; PST: Peri-Stimulus Time; QC: Quality Control; μV: Microvolts

| Step | Parameter | Value |
| --- | --- | --- |
| Oddball paradigm | Inter-trial interval | 500 ms |
|  | Tone length | 70 ms |
|  | Visual event presentation time | randomly 50-250 PST |
|  | Total number of trials | 1800 |
| Filtering and downsampling | Sampling rate | 2048 Hz |
|  | Downsampling rate | 256 Hz |
|  | Low-pass filter cut-off frequency | 30 Hz |
|  | High-pass filter cut-off frequency | .5 Hz |
|  | Filter type | 5th order Butterworth |
| **ICA and artefact rejection** | Components | 30 |
|  | Correlation threshold for removing eye ICA components | .7 |
|  | ICLabel probability threshold | 75% |
|  | ICA component kurtosis threshold | 15 |
| | Artefact rejection amplitude threshold | $\pm 80 \mu\text{V}$ |
| **Epoching** | Epoch length | 550 ms |
|  | Time interval | -100 ms to 450 ms PST |
| **Averaging** | Method | SPM-Robust |
|  | Binning threshold for model-based ERPs | 20% |
| **Statistical analysis** | Image grid | 32×32 |
|  | Smoothing kernel | 16×16 mm |
|  | Time window | 100-400 PST |
|  | F-test significance threshold | 11.34 |
|  | T-test significance threshold | 3.15 |
| **QC** | Number of artefact-free trials | >1000 |
|  | Number of bad channels | <5 |
|  | Number of removed ICA components | <26 |
|  | Number of total artifact probability among retained ICA components | <25% |
|  | Visual task hit rate | >50% |

### EEG Acquisition and Preprocessing

Continuous 64-channel EEG and EOG were acquired using a BioSemi ActiveTwo system. Pre-processing, quality control (QC), and statistical analyses were conducted via SPM12 (v7771) in MATLAB (R2023b), utilizing EEGLab (v2025.1.0) and ICLabel (v1.7) for automated ICA-based artefact removal (57–59). Following filtering, average-referencing, and epoching, overall and phase-specific MMN difference waveforms were generated by subtracting standard from deviant ERPs. Full pipeline specifications are detailed in Table 1 and Supplementary Material S2–S4.

### Computational Modelling: Hierarchical Gaussian Filter

Visualized in Figure 1.B is the 3-level HGF structure used in this study. Because the oddball paradigm does not require behavioural responses, participants were modelled as surprise-minimising Bayesian observers, consistent with prior applications of the HGF to MMN paradigms (14,60). Via variational Bayesian inversion, trial-by-trial computational quantities were derived using the HGF (version 6) (37,61). Finally, perceptual parameters were optimised using the tapas_ehgf_binary function (TAPAS v4.0.0; https://github.com/ComputationalPsychiatry) (62).

The following regressors were extracted for EEG analysis: ε_2_ (low-level sensory pwPE); ε_3_ (volatility-level pwPE); δ_1_ (unsigned low-level PE); δ_2_ (unsigned high-level PE); ψ_2_ (level-2 posterior uncertainty); ψ_3_ (level-3 posterior uncertainty). The belief updating (Δμ) at level i and trial k is done via the following equation:

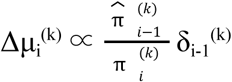

Here, δ_i-1_^(k)^ represents the precision-weighted prediction error used to update the belief at level i. This error is generated by the discrepancy between the expected and actual outcomes at the subordinate level (i-1). When relayed to the upper level, the prediction error is scaled by a precision ratio: the update is directly proportional to the precision of the lower-level prediction and inversely proportional to the precision of the current belief at level i. The precision ratio can adjust belief updating based on the environment’s volatility or stability. More details on the HGF model is included in the supplementary Material Section S5.

Consistent with Weber, Diaconescu et al. (60) and Charlton et al. (14), regressors entered the general linear model (GLM) without mutual orthogonalization, preserving their natural correlational structure. In the model-based ERP analysis, standards (low condition) were defined as trials in the bottom 20% of each regressor, and deviants (high condition) as those in the top 20%. Trajectories of the ideal observer’s precisions, prediction errors and pwPEs are presented in Figure 1.C.

### Statistical Analysis

Spatiotemporal volumetric images were generated for each condition by projecting electrode voltages onto a standardized grid and using linear interpolation to create a continuous spatial field. These sequential scalp maps were stacked across peristimulus time and smoothed with a Gaussian kernel to meet random field theory assumptions (59,63). Finally, analyses were restricted to 100-400 Peri-Stimulus Time (PST) to capture relevant MMN and P300 activity. All second-level analyses were conducted in SPM12 across time × scalp channel space, with family-wise error (FWE)-corrected cluster-level inference. Age and sex were entered as nuisance covariates in all group-level contrasts.

## Results

### Sample Characteristics and Bifactor Analysis

Group characteristics for the QC-passed EEG subset (N=131; PSS-=66; PSS+=65) are summarized in Table 2. They differ significantly in age (T=2.96, p<0.01) and sex assigned at birth (χ²=9.31, p=.009) with PSS+ youth comprising a larger proportion of females and gender-diverse individuals. Ethnicity did not differ between groups. PRIME-R total scores were markedly higher in PSS+ youth (T=−11.40, p<0.0001) with significantly lower functioning. PSS+ also had significantly higher levels of anxiety, mood and trauma-related disorders.

**Table 2.** Sample characteristics. Continuous variables are reported as mean (std)

| Characteristics |  | PSS-, N=66 | PSS+, N=65 | Test Statistics |
| --- | --- | --- | --- | --- |
| Age* | | 16.22 (3.39) | 17.91 (3.15) | $T = 2.96$<br>$p = .003$ |
| Sex* | Female | 37 | 51 | $\chi^2 = 9.31$<br>$p = .009$ |
|  | Male | 29 | 13 |  |
|  | PNA<sup>2</sup> | 0 | 1 |  |
| Gender | Boy or Man | 25 | 14 | $\chi^2 = 5.31$<br>$p = .065$ |
|  | Girl or Woman | 28 | 34 |  |
|  | Non-binary or another gender identity | 7 | 13 |  |
|  | Other<sup>3</sup> | 6 | 4 |  |
| **Ethnicity** | White | 46 | 34 | $\chi^2 = 7.80$<br>$p = .099$ |
|  | Black | 4 | 2 |  |
|  | Asian | 2 | 6 |  |
|  | Mixed | 12 | 16 |  |
|  | Other | 2 | 7 |  |
| **PRIME-R total score** | | 14.76 (12.52) | 39.54 (12.36) | $T = -11.40$<br>$p < .001$ |
| **Global functioning** | Role<sup>*4</sup> | 6.95 (2.06) | 5.96 (2.58) | $T(124) = 2.38$<br>$p = .02$ |
| | Social<sup>*5</sup> | 7.56 (1.24) | 7.01 (1.45) | $T(126) = 2.31$<br>$p = .02$ |
| **Clinical diagnosis** | Anxiety disorders* | 37 | 49 | $\chi^2 = 6.84$<br>$p = .009$ |
| | Any mood conditions* | 7 | 23 | $\chi^2 = 12.12$<br>$p < .001$ |
| | Any neurodevelopmental disorders | 25 | 14 | $\chi^2 = 3.74$<br>$p = .053$ |
| | Any trauma and stress-related disorders* | 5 | 17 | $\chi^2 = 8.58$<br>$p = .003$ |
| | Any Schizophrenia and psychotic disorders | 1 | 4 | $\chi^2 = 2.02$<br>$p = .15$ |

<sup>1</sup> Asterisks indicate significant group differences ( $p_{\text{value}} < 0.05$ ).
<sup>2</sup> PNA: Prefer Not to Answer.
<sup>3</sup> Other includes Prefer not to answer, unknown, skipped response and missing.
<sup>4</sup> Five subjects had missing GF:Role scores.
<sup>5</sup> Three subjects had missing GF:Social scores.

The model identified a robust general factor with substantial loadings across all items (λ = 0.48–0.71; see Figure 2.B). Two orthogonal specific factors were retained: Specific_1 and Specific 2 (see Figure 2.C,D). Specific_1 captures “Unusual thought content, delusional mood” (dominant items: q9=0.50, q12=0.46, q10=0.44, q5=0.43), reflecting the attenuated psychosis syndrome core. Specific_1 factor reflects the top items scored by the participants (compare Figure 2.A and loadings in Figure 2.C). Specific 2 solely represents the “Grandiosity/superstitions” dimension (dominant items: q2=0.49, q4=0.29, q8=0.42), reflecting positively valenced overvalued ideation. However, this sub-dimension exhibited sub-optimal construct replicability (H-index = 0.589). The rest of the fitting metrics for the factors are reported in section S1.5 of the Supplementary Material.

**Figure 2.**
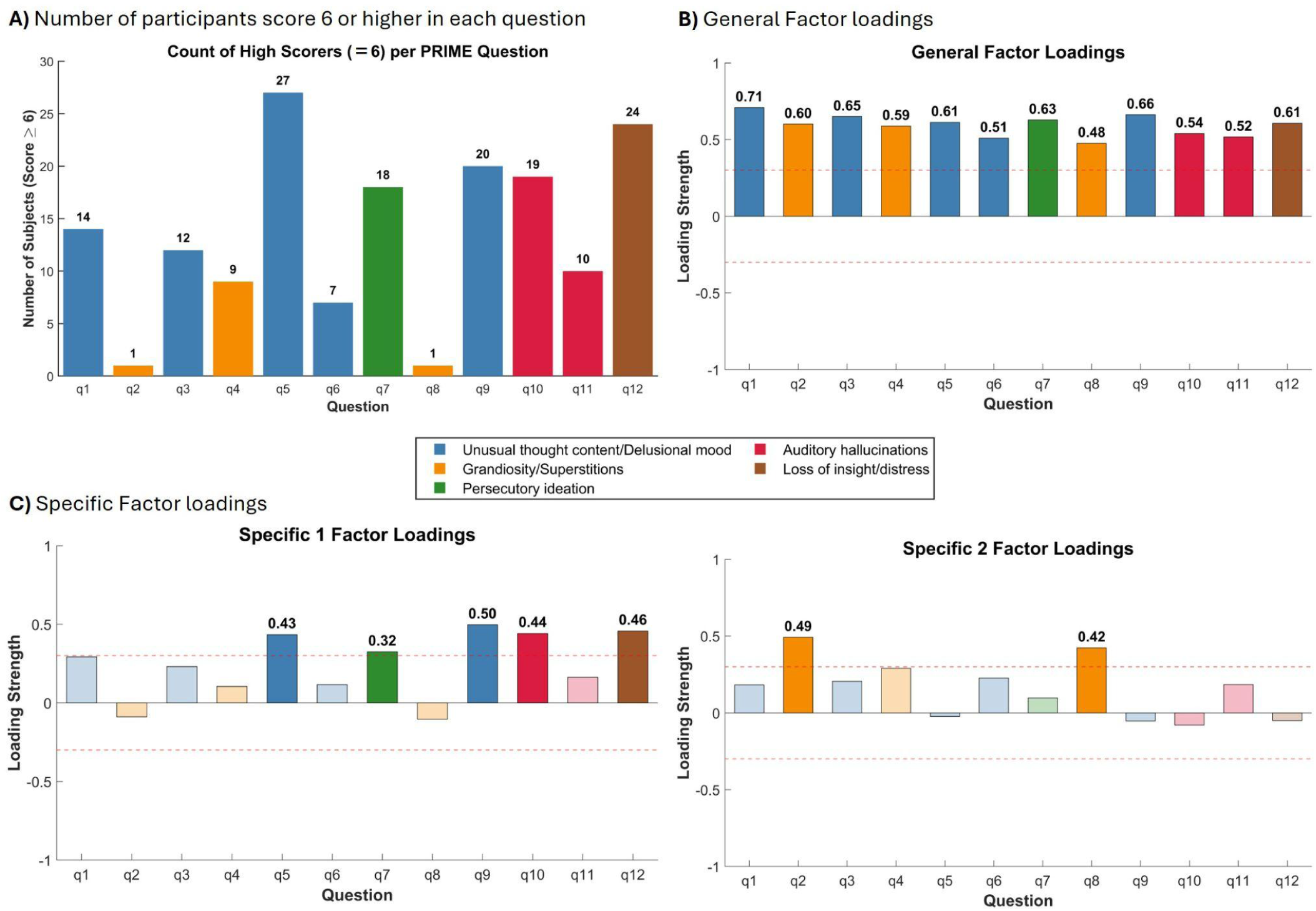
General and specific factor loadings for the 12-item PRIME-R. **A)** The high scorers (score = 6) are counted and shown per PRIME-R items. **B-C)** Loading strengths are shown for the General Factor and two Specific Factors (left, right). Symptom subscales are defined as Unusual Thought Content/delusional mood (items 1, 3, 5, 6 and 9), Suspiciousness (item 7), Grandiosity/Superstitions (items 2, 4 and 8), Persecutory Ideation (item 7) and Auditory Hallucinations (items 10–11) and Loss of insight/distress (item 12). The red dotted line indicates the ±0.3 loading threshold. Significant loadings (≥ 0.3) are annotated and shown in full opacity, whereas sub-threshold loadings are faded. The General Factor reflects a strong common variance across all 12 items. Specific 1 highlights residual variance in unusual thought content, persecutory ideation, and auditory hallucinations and match the top items across the participants, while Specific 2 distinctly captures grandiosity and superstitions.

### Paradigm Validation

#### Main Effect of Oddball MMN

A robust main effect of the oddball MMN (deviant−standard) was observed across the full QC-passed sample peaking at 188 and 297 ms (see Table 3 for more details). Multiple FWE-corrected clusters spanning fronto-central and bilateral temporal channels were identified, consistent with canonical MMN topology (16,59). Figure 3 demonstrates scalp topographies and frontocentral midline electrodes’ ERPs and difference waveform for oddball MMN and MMN in stable and volatile phases.

**Figure 3.**
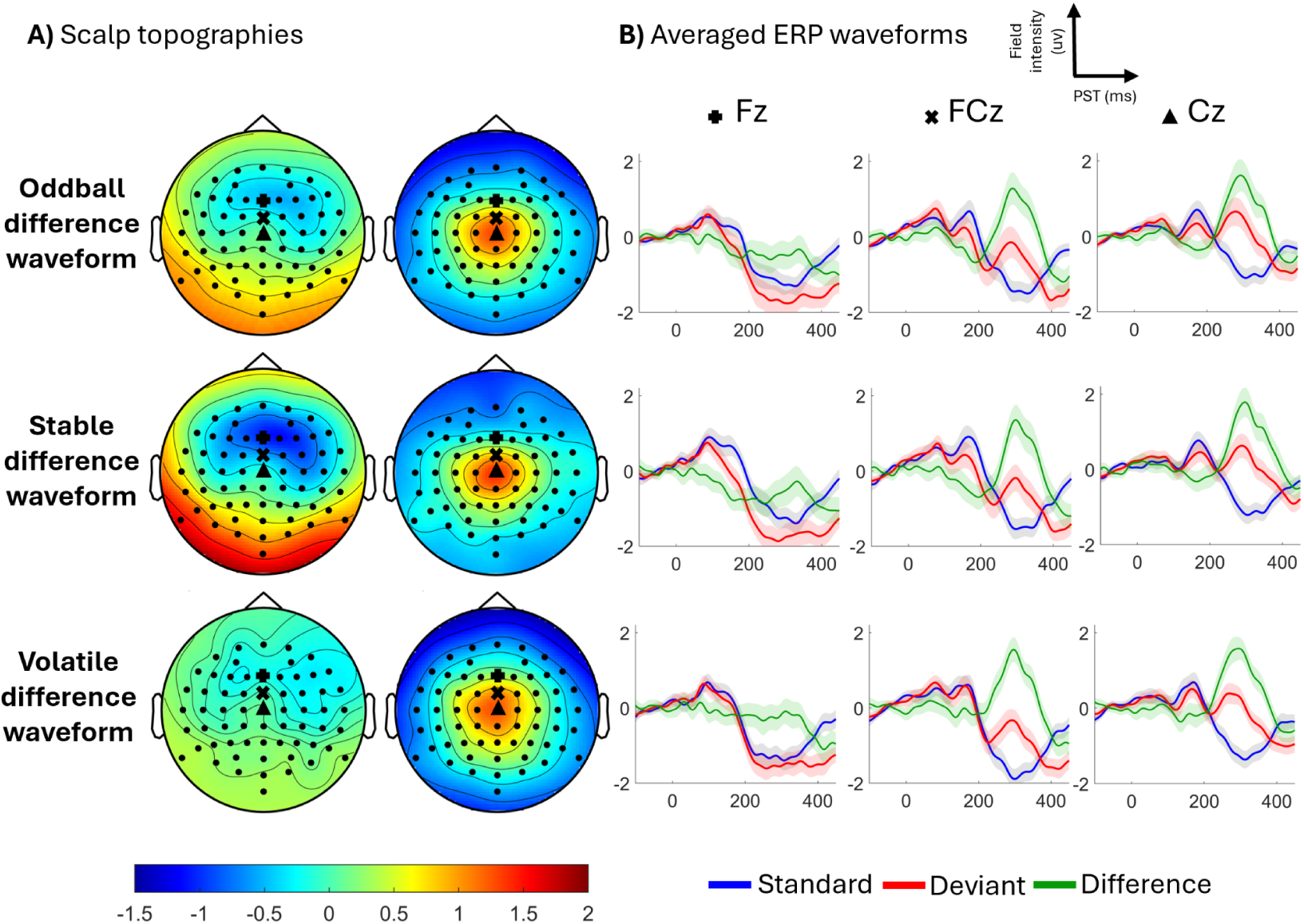
Grand average ERPs and scalp topographies for standard, deviant, and difference waveforms. **(A)** Topographical maps of the difference waveform for the overall oddball, stable, and volatile conditions, capturing early frontocentral negativity and later central positivity. Midline electrodes (Fz, FCz, Cz) are marked. **(B)** Stimulus-locked ERP waveforms at Fz, FCz, and Cz. Responses to standard (blue) and deviant (red) tones are plotted with the resultant difference waveform (green). Shaded areas represent the standard error of the mean. Rows correspond to the overall sequence, stable phases, and volatile phases, respectively.

**Table 3.** Summary of key results. For each regressor, the top four significant distinct clusters are listed. Cells with significant peak-level or cluster level FWE-corrected p-values are highlighted. Clus. Num: cluster number; Z_E_: equivalent z-score; k_E_: cluster size; peak p_FWE_: peak-level family-wise error corrected p-value; MMN: mismatch negativity; GF: Global Functioning;

| | Regressor | Clus. Num. | F/T value | $Z_E$ | Peak -level $p_{\text{FWE}}$ | $k_E$ | Peak Coord. (mm, mm, ms) | Time Range [$t_{\text{low}}$, $t_{\text{high}}$] |
| --- | --- | --- | --- | --- | --- | --- | --- | --- |
| Paradigm Validation | Main oddball MMN effect | 1 | $F_{(1,130)}=141.43$ | Inf | <.001 | 12496 | (0, -9, 297) | [109, 398] |
| | | 2 | $F_{(1,130)}=58.09$ | 6.82 | <.001 | 15997 | (34, 2, 188) | [117, 398] |
| | Stable MMN < Volatile MMN | 1 | $T_{(130)}=6.07$ | 5.68 | <.001 | 3113 | (-47, -73, 230) | [141, 273] |
| | Stable MMN > Volatile MMN | 1 | $T_{(130)}=5.49$ | 5.20 | <.001 | 2400 | (26, 18 238) | [121, 262] |
| | $\epsilon_2$ main effect | 1 | $F_{(1,130)}=201.77$ | Inf | <.001 | 14896 | (-42, -73, 184) | [102, 398] |
| | | 2 | $F_{(1,130)}=191.48$ | Inf | <.001 | 18750 | (38, 8, 176) | [113, 398] |
| | $\epsilon_3$ main effect | 1 | $F_{(1,130)}=266.55$ | Inf | <.001 | 15866 | (-47, -68, 195) | [102, 398] |
| | | 2 | $F_{(1,130)}=212.12$ | Inf | <.001 | 21740 | (38, 2, 180) | [102, 398] |
| | $\delta_1$ main effect | 1 | $F_{(1,130)}=201.36$ | Inf | <.001 | 15607 | (30, 13, 176) | [121, 398] |
| | | 2 | $F_{(1,130)}=195.24$ | Inf | <.001 | 14360 | (-42, -73, 188) | [105, 398] |
| | | 3 | $F_{(1,130)}=23.72$ | 4.51 | .006 | 569 | (47, -68, 293) | [277, 316] |
| | $\delta_2$ main effect | 1 | $F_{(1,130)}=158.52$ | Inf | <.001 | 16287 | (-21, -84, 176) | [109, 398] |
| | | 2 | $F_{(1,130)}=131.69$ | Inf | <.001 | 19394 | (38, 24, 176) | [113, 398] |
| | $\psi_2$ main effect | 1 | $F_{(1,130)}=23.23$ | 4.47 | .008 | 384 | (-13, -19, 340) | [324, 371] |
| | | 2 | $F_{(1,130)}=19.32$ | 4.08 | .037 | 361 | (8, -3, 293) | [273, 309] |
| | | 3 | $F_{(1,130)}=19.13$ | 4.06 | .040 | 602 | (8, 61, 207) | [195, 230] |
| | $\psi_3$ main effect | 1 | $F_{(1,130)}=29.04$ | 4.98 | .001 | 966 | (-4, -3, 301) | [281, 367] |
| | | 2 | $F_{(1,130)}=21.82$ | 4.33 | .014 | 1427 | (30, 40, 207) | [191, 227] |
| | | 3 | $F_{(1,130)}=20.62$ | 4.21 | .022 | 660 | (42, -52, 223) | [172, 238] |
| | Volatile MMN - GF: Social | 1 | $T_{(124)}=4.26$ | 4.11 | .029 | 496 | (-51, -41, 301) | [254, 316] |
| Age | Oddball MMN | 1 | $F_{(1,126)}=15.58$ | 3.65 | .15 | 412 | (0, -52, 324) | [297, 336] |
| | $\delta_1$ | 1 | $F_{(1,126)}=27.2$ | 4.82 | .002 | 853 | (38, -52, 227) | [207, 254] |
| | | 2 | $F_{(1,126)}=18.49$ | 3.98 | .053<sup>†</sup> | 486 | (13, 50, 227) | [215, 250] |
| | $\delta_2$ | 1 | $F_{(1,126)}=24.86$ | 4.61 | .005 | 1034 | (-8, -62, 324) | [297, 355] |
| | | 2 | $F_{(1,126)}=23.93$ | 4.53 | .007 | 918 | (13, 50, 328) | [297, 352] |
| | | 3 | $F_{(1,126)}=16.62$ | 3.77 | .111<sup>†</sup> | 443 | (8, 45, 230) | [219, 242] |
| | | 4 | $F_{(1,126)}=16.25$ | 3.73 | .127 | 337 | (4, -52, 234) | [223, 242] |
| Categorical | $\epsilon_3$, PSS+ < PSS- | 1 | $T_{(126)}=4.30$ | 4.14 | .024 | 379 | (-21, -3, 180) | [164, 195] |
| | $\epsilon_3$, PSS+ > PSS- | 1 | $T_{(126)}=3.46$ | 3.38 | .30 | 106 | (-21, -84, 184) | [152, 188] |
| Dimensional | $\delta_2$, General factor | 1 | $T_{(126)}=4.02$ | 3.89 | .065<sup>†</sup> | 471 | (-8, 8, 137) | [125, 148] |
| | $\psi_2$, Specific_1 factor | 1 | $T_{(124)}=4.02$ | 3.89 | .070 | 332 | (-4, -46, 387) | [367, 398] |

<sup>†</sup> Only cluster-level significant ( $p_{\text{FWE-clus}} < .05$ )

#### Stable versus Volatile Phase Comparison

The MMN amplitude significantly reduced in the volatile-phase [T(130)=6.07; p_FWE-peak_<.0001; peaks: (−47,−73, 230ms)] (Figure 4.A-C), replicating prior demonstrations of context-dependent predictive coding (14,15). The volatile-phase MMN also exceeded the stable-phase in a distinct cluster [T(130)=5.49; p_FWE-peak_<.0001; peak: (26, 18, 238ms)] (Figure 4.D-F), indicating bidirectional phase modulation of auditory deviance detection.

**Figure 4.**
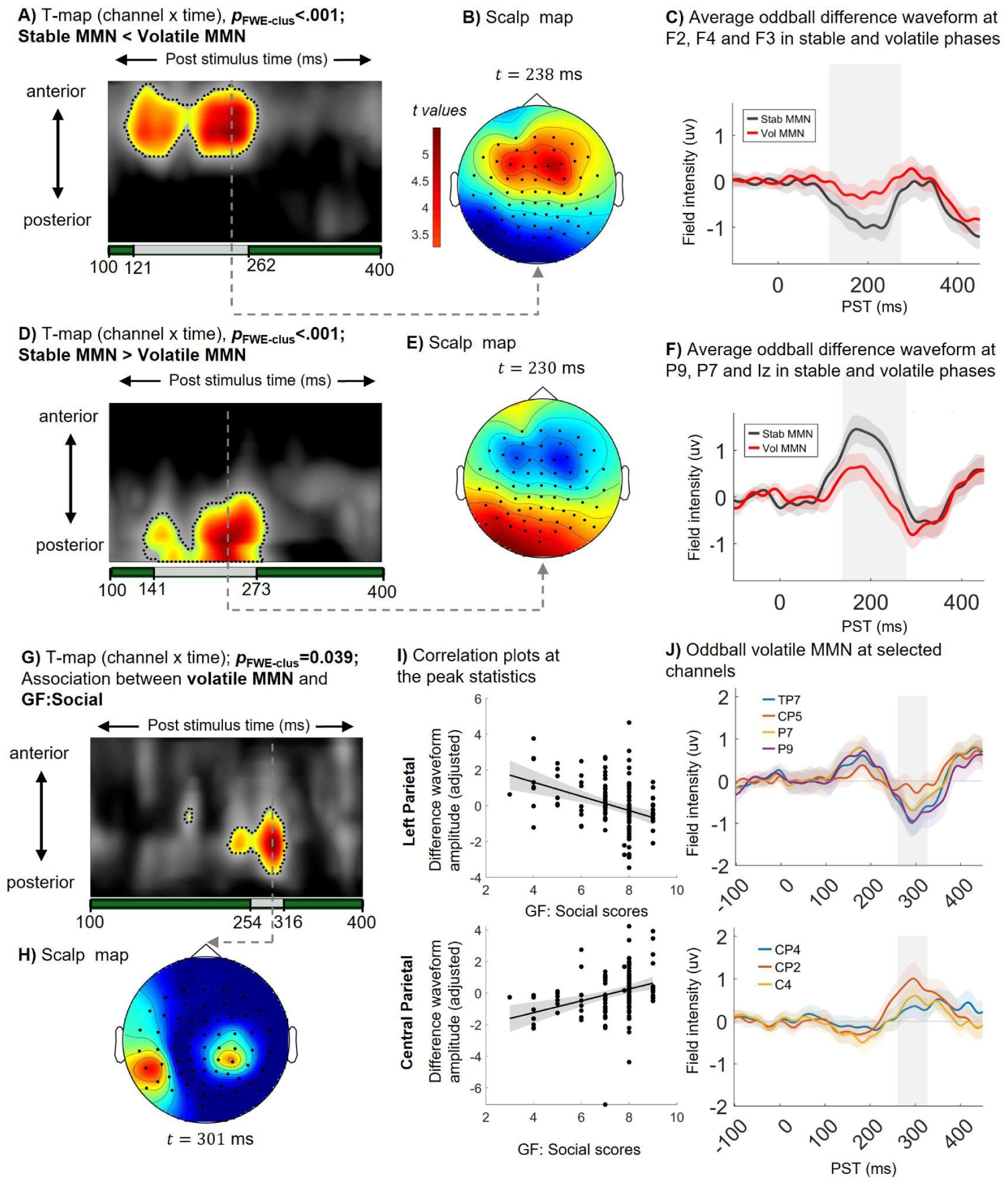
Stable vs. volatile phase comparison and association with social functioning. (A-C) Significant anterior cluster (p_FWE-clus_ < .001) showing a reduced (more negative) MMN during stable compared to volatile phases between 121–262 ms, visualized via T-map, scalp topography at 238 ms, and average waveforms at F2, F3, and F4. **(D-F)** Significant posterior cluster (p_FWE-clus_ < .001) showing enhanced MMN for the stable phase from 141–273 ms, shown with topography at 230 ms and waveforms at P9, P7, and Iz. **(G-J)** T-map reveals a significant association (p_FWE-clus_ = .039) between the volatile difference waveform and GF:Social scores in a later time window (254–316 ms). Scatter plots show the directionality of the correlations at peak statistics, with corresponding waveforms plotted for the relevant channels. MMN: mismatch negativity; PST: peri-stimulus time; FWE: family wise error. GF: global functioning.

#### Age as the Dominant Modulator of Sensory-Level Predictive Processing

Age was the dominant source of EEG variance across sensory-level regressors, driving significant effects across the δ_1_ [F(1,126) = 27.20; p_FWE-peak_ = .002; k_E_ = 853; p_FWE-clus_ = .006; peak voxel: (38, −52, 227 ms)] peaking at an earlier time window spanning 207-250 ms (see Figure 5.A-D) and oddball MMN [F(1,126) = 15.58; p_FWE-peak_ .147; k_E_ = 412; p_FWE-clus_ = .044; peak voxel: (0, −52, 324 ms)] and δ_2_ [F(1,126) = 24.86; p_FWE-peak_ .005; k_E_ = 1034; p_FWE-clus_ = .003; peak voxel: (-8, −62, 324 ms)] (see Figure 5.E-H) peaking at a later window including 300-340 ms. We observed a characteristic frontal-positive/parietal-inverse pattern.

**Figure 5.**
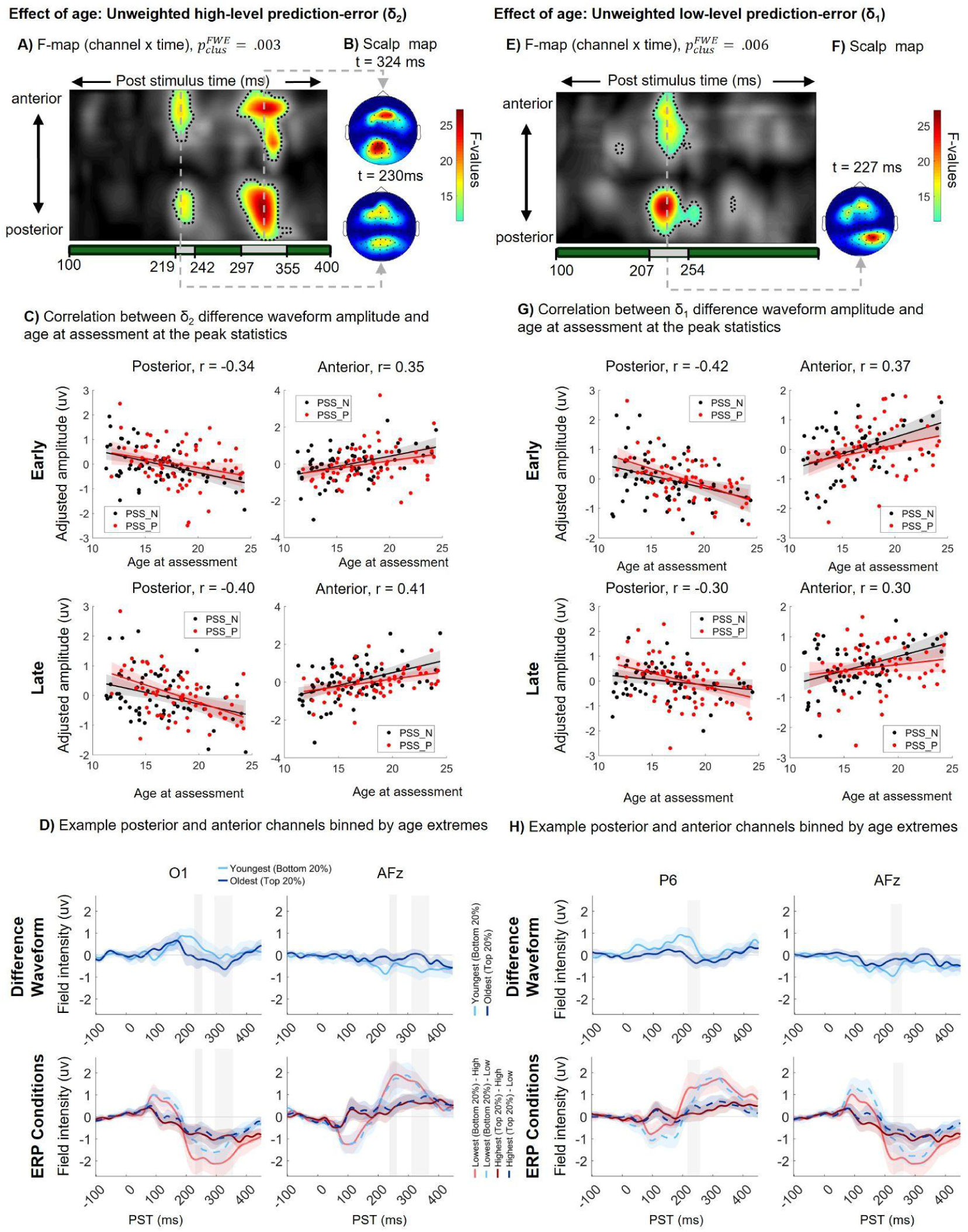
Effect of age on unweighted prediction errors (δ_1_ and δ_2_). Analysis of spatiotemporal relationships between unweighted high-level prediction-error (δ_2_) and low-level prediction-error (δ_1_) waveforms and subject age. **(A-D)** Effect of age on unweighted high-level prediction-error (δ_2_): **(A)** Channel-x-time F-maps highlighting dynamic relationships between δ_2_ and age, with significant clusters outlined (p_FWE-clus_ = .003). Shaded regions below show significant time windows. **(B)** Scalp topographies at key statistical peak times (t=230 ms and t=324 ms). **(C)** Correlation plots showing early/late and anterior/posterior relationships at peak statistics. Plots for “adjusted amplitude vs. age” include best-fit lines and significance thresholds. **(D)** Example workflows for representative posterior (O1) and anterior (AFz) channels, binned by age extremes (Youngest bottom 20%, Oldest top 20%). Separate plots show Difference Waveform versus complete ERP Conditions (Standard, Deviant). (E-H) Effect of age on unweighted low-level prediction-error (δ_1_): **(E-H)** are structured in parallel to (A-D), detailing F-maps, topographies (t=227 ms), correlation plots, and binned age waveforms for δ_1_ prediction-error analyses, with significant clusters enclosed by dashed lines. All waveforms and correlation plots use adjusted scores for optimized visualization of the effects.

#### Categorical PSS Marker: ε_3_ (Volatility-Level pwPE)

The ε_3_ regressor yielded significant main effects in both PSS+ [F(1, 64)=187.88; p_FWE-peak_<0.0001; k=10,866; peak: (−34, −78, 184 ms)] and PSS– [F(1, 65)=106.77; p_FWE-peak_<.0001; k=6,404; peak: (−47,−68, 195 ms)] subgroups, confirming robust volatility-level pwPE encoding in both groups. The PSS+<PSS– contrast yielded a significant peak after controlling for age and sex [T(126)=4.30; p_FWE-peak_=.024; k_E_=379; p_FWE-clus_=.063; peak: (−21, −3, 180 ms)], indicating reduced ε_3_ modulation (more negative) in PSS+ youth at central-frontal channels (see Figure 6.A-C). This is the only model-based contrast that is not confounded by age or sex, which are entered as covariates of no interest (age: F(1,126)=14.79, p_FWE-peak_=.186, peak: (34, -52, 227 ms); sex: F(1,126)=13.15, p_FWE-peak_=.32, peak: (-64, 2, 203 ms)). The PSS+>PSS– contrast did not yield a significant FWE-corrected cluster after controlling for age and sex [T(126)=3.46; p_FWE-clus_=0.21; k_E_=106; p_FWE-peak_=0.26; peak: (−21,−84, 184 ms)] (see Figure 6.D-F).

**Figure 6.**
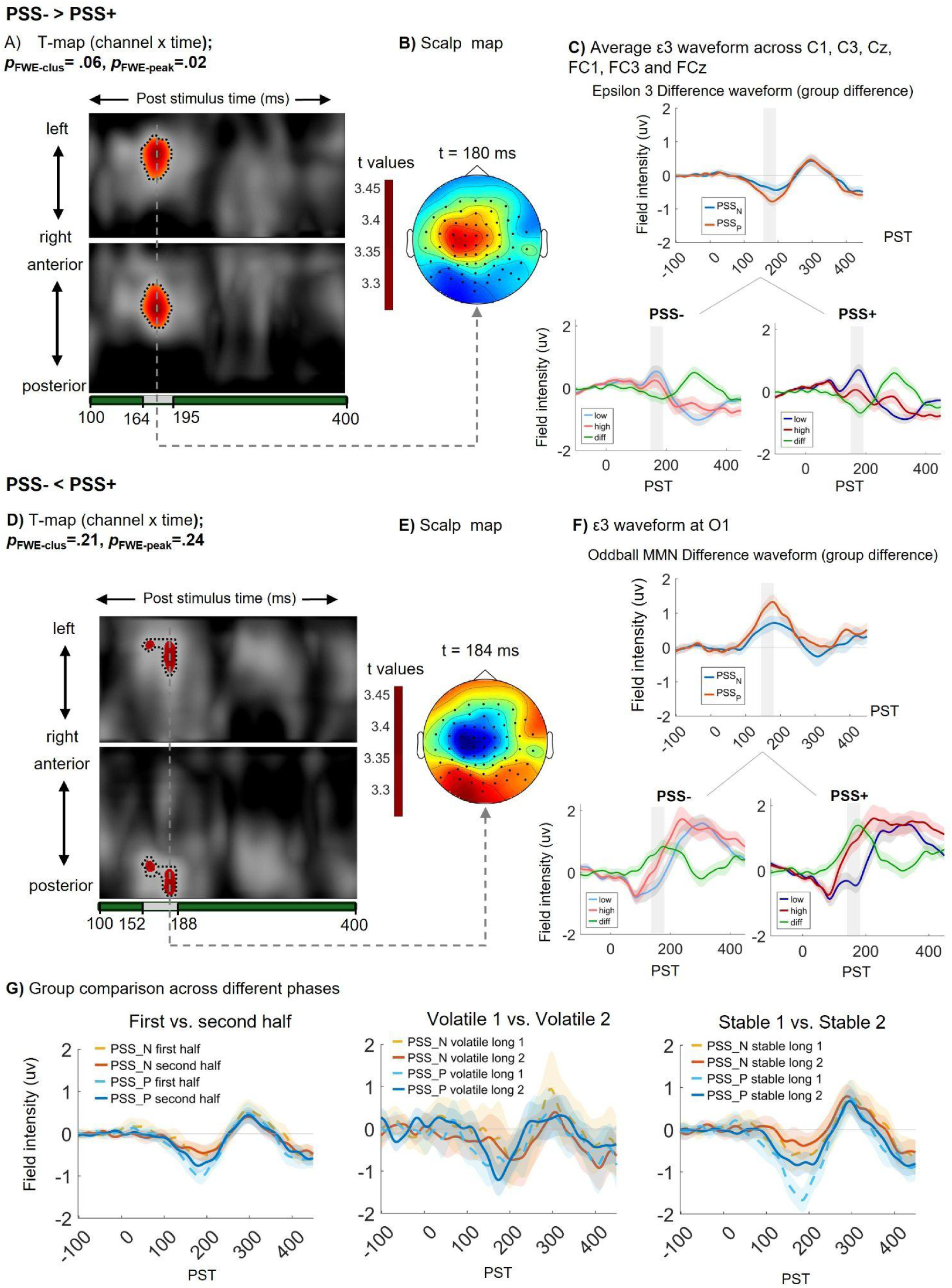
Categorical PSS Marker: ε_3_ (Volatility-Level pwPE) (PSS- vs. PSS+). **(Top)** PSS- > PSS+ Contrast: **(A)** Statistical T-map shows a trend-level cluster (p_FWE-clus_ = .06) from 164–195 ms. **(B)** Scalp map at t = 180 ms shows a central positivity. **(C)** Top right: Direct group comparison of the ε_3_ difference waveform averaged across a frontocentral cluster (C1, C3, Cz, FC1, FC3, FCz). Bottom right: Condition-specific ERPs plotted separately by group. (Bottom) PSS- < PSS+ Contrast: **(D)** T-map reveals a cluster from 152–188 ms (p_FWE-clus_ = .21). **(E)** Corresponding scalp topography at t = 184 ms. **(F)** Top right: Group comparison of the ε_3_ difference waveform at electrode O1. Bottom right: Within-group condition ERPs. Grey shaded bars indicate the time windows of the respective statistical clusters. **(G)** Temporal trajectories of the ε_3_ response across the different phases of the paradigm.

The PSS+<PSS– contrast remained significant after controlling for the total number of comorbidities [T(125)=4.31; p_FWE-peak_=.024; k_E_=360; p_FWE-clus_=.068; peak: (−21, 2, 180 ms)]. Moreover, we assessed the correlation between ε_3_ and the top three PRIME-R individual items that load most heavily onto the Specific_1 factor. Question 9—which assesses unusual thought content, particularly the experience of ‘mind tricks’—demonstrated a significant correlation, yielding a voxel cluster that overlapped with the main group effect [F(1,126) = 18.85; p_FWE-peak_ = 0.047; k_E_ = 284; p_FWE-clus_ = 0.077; peak: (-26, 2, 184 ms)].

ε_3_ did not show a significant dose-response relationship with PRIME-R total score nor with either bifactor subscale. The ε_3_ signal therefore constitutes a categorical neural marker but does not track where individuals fall on a continuous symptom severity continuum. (11)

Finally, Figure 6.G depicts the ε_3_ trajectories across the temporal unfolding of the paradigm at different phases. The enhanced frontocentral negativity in the PSS+ group was driven primarily by trials within the initial stable phase (stable_1) while during the second stable phase (stable_2), the ε_3_ response in the PSS+ group diminished to levels comparable to the PSS– group.

### Dimensional PSS Severity Marker: δ_2_ (Unweighted Volatility-Level PE)

δ_2_ was the only regressor showing a significant inverse association with the General PSS factor [T=4.02; p_FWE-peak_=.065; k_E_=471; p_FWE-clus_=0.04; peak: (-8, 8, 137)]: higher transdiagnostic PSS burden was associated with reduced unweighted volatility-level PE amplitude (see Figure 7.A-D). The δ_2_ group contrast (PSS+ vs. PSS–) did not reach FWE-corrected significance, indicating δ_2_ does not function as a categorical group marker.

**Figure 7.**
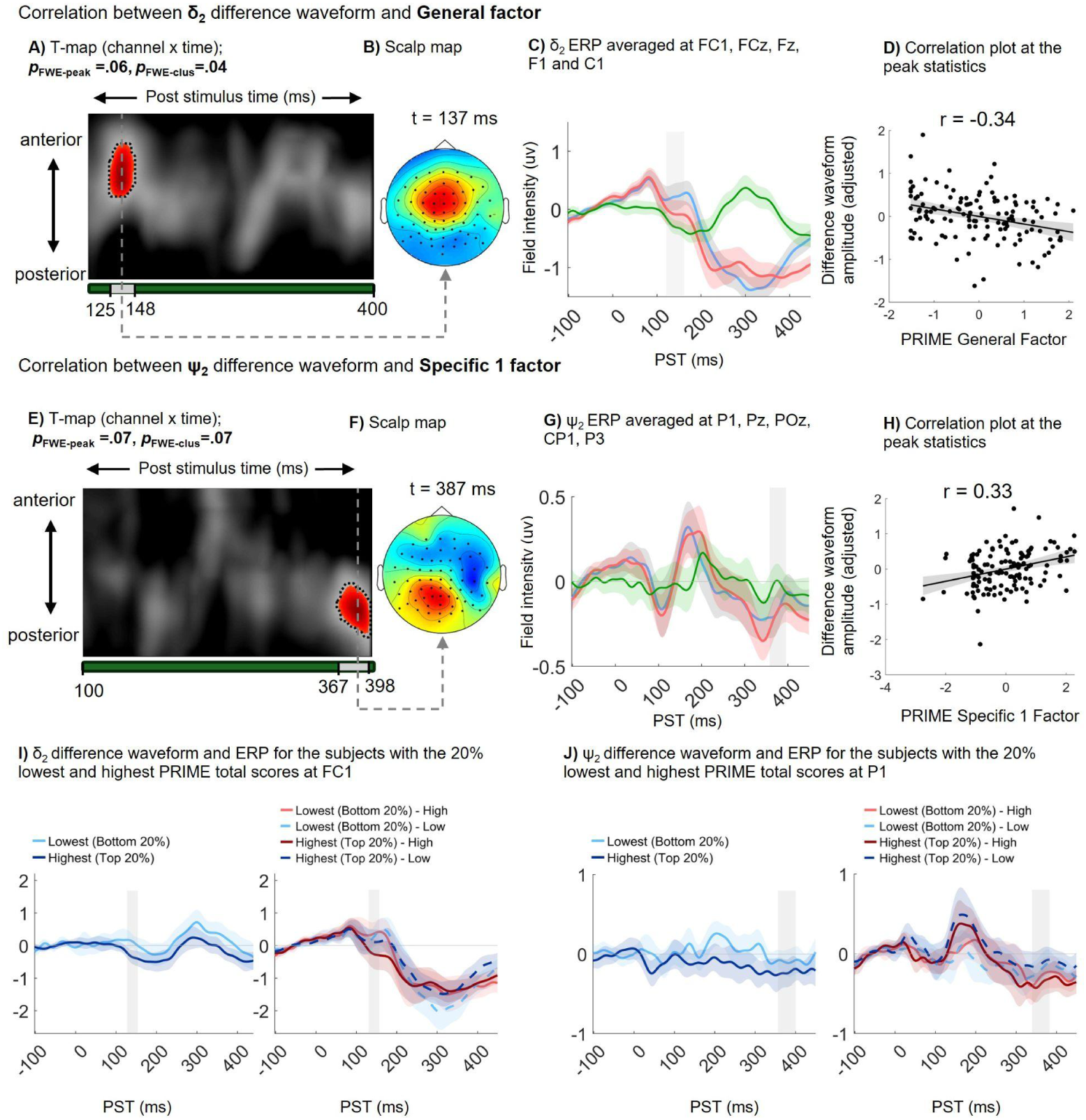
Dimensional PSS Severity Marker: δ_2_ (Unweighted Volatility-Level PE). **(Top row, A-D):** The PRIME General Factor shows a significant negative correlation (Pearson’s r = -0.34, p_FWE-clus_ = .04) with the δ_2_ difference waveform in an early anterior cluster (125–148 ms). Shown via T-map, scalp topography (t = 137 ms), averaged waveforms at frontocentral electrodes, and scatter plot at peak statistics. **(Middle row, E-H):** The PRIME Specific_1 Factor exhibits a trend-level positive correlation (Pearson’s r = 0.33, p_FWE-clus_ = .07) with the ψ_2_ waveform in a later posterior cluster (367–398 ms), illustrated with corresponding T-map, topography (t = 387 ms), posterior waveforms, and scatter plot. **(Bottom row, I-J):** Visualization of these effects comparing subjects with the 20% lowest vs. highest PRIME total scores, displaying both the overall difference waveforms and the separated condition ERPs at representative anterior (FC1) and posterior (P1) channels.

The ψ_2_ regressor (level-2 posterior uncertainty) showed a trend-level positive association with the Specifc 1 PSS factor [T(124)=4.02; p_FWE-peak_=.07; k_E_=332; p_FWE-clus_=.066; peak: (−4,−46, 387 ms)] (see Figure 7.E-H). No significant dimensional association was found between Specific 2 factor, that represents grandiosity, and difference waveforms.

### Social Functioning Specificity: Volatile-Phase MMN and GF:Social

The volatile-phase MMN showed a significant negative association with GF:Social at temporal channels around 300 ms [T(124)=4.26; p_FWE-peak_=.029; k_E_=496; p_FWE-clus_=.039; peak: (-51, −41, 301 ms)]. Visual inspection of the difference waveform at the peak channels show a negative-going waveform (see Figure 4.I). No equivalent association emerged in the stable phase. No significant association with GF:Role was observed in either phase.

This directly replicates the finding reported by Charlton et al. (14) in non-clinical controls using the identical paradigm: volatile-phase MMN positively associated with GF:Social at frontocentral channels (Fz), peaking at 344 ms [T(41)=4.8; pFWE=0.036]. Although the peak electrode (temporal in TAY vs. frontocentral in Charlton et al. (14)) and timing (300 ms vs. 344 ms) differ slightly, both findings fall within the P300 window and share the same functional specificity of the volatile context. The cross-sample directional consistency supports the robustness of this association.

### Summary of Key Results

Table 3 provides a structured overview of primary statistical results across all contrasts and model-based regressors.

## Discussion

The present study applied a hierarchical Bayesian framework to EEG recordings during a stable/volatile auditory oddball paradigm in 131 youth seeking mental health care for a variety of mental health presentations at a tertiary-level psychiatric hospital setting. Our findings of increased volatility-level pwPEs (ε_3_) reveal that disruption of predictive coding in PSS is specific to the volatility level of the auditory hierarchy. We discuss these in turn, followed by their collective implications.

### Age as the Dominant Modulator of Sensory-Level Predictive Processing

Age was the dominant source of EEG variance across the sensory-level regressors (oddball ERP and δ₁) and higher-order volatility regressors (δ₂), manifesting as a characteristic frontal-positive/parietal-inverse pattern at early and late time windows spanning 207–254 ms and 300-336 ms PST, respectively. Specifically, the significant age-related variance in the oddball difference response emerged outside the canonical MMN window, aligning instead with the P300 interval, a component reflecting higher-order novelty processing, context updating, and attention-orienting mechanisms (64–66). Consistent with the meta-analytic findings of van Dinteren et al. (65), P300 amplitude increases and latency decreases through late adolescence, reaching a peak around age 20 before beginning a gradual decline. Additionally, the δ₁-binned difference waveform exhibited age-related reductions in both latency and amplitude, manifest as diminished frontal negativity and attenuated posterior positivity.

Unlike the P300, the MMN has traditionally been regarded as developmentally stable, maturing mainly through modest latency reductions rather than amplitude change. The descriptive literature is consistent on this contrast: across childhood and adolescence the P3a/P300 changes markedly in amplitude and latency, whereas the MMN follows a shallow inverted-U from infancy through the early school years into adulthood (67–69). To our knowledge, the present study is the first to apply HGF modelling to age effects on the auditory mismatch response. The maturational change we detect lies in the early sensory prediction error (δ₁), within the canonical MMN window. Because conventional MMN averaging collapses the whole predictive-coding hierarchy into a single difference waveform, this refinement might be invisible to it—plausibly why the averaged MMN has so often appeared “stable.”

Tracking adolescent synaptic pruning, progressive myelination, and the formation of long-range connections (70–72), these trajectories might suggest a hierarchically segregated refinement of belief-updating: basic deviance detection (δ₁) demands fewer resources as lower-order cortices mature, whereas the capacity to track environmental volatility (δ_2_) develops slower alongside higher-order prefrontal-parietal networks (73–75). Crucially, their precision-weighted counterparts (ε_2_ and ε_3_) showed no significant age effects. This implies that while the raw magnitude of prediction errors undergoes marked maturational change, the relative confidence weighting these errors against prior beliefs—the core inferential operation of hierarchical Bayesian learning (38)—remains developmentally stable across the 11–24 year age range.

### Volatility Precision-Weighted PE as a Categorical Neural Marker of PSS

The primary finding of this study is a statistically significant PSS group difference in the volatility-level precision-weighted prediction error regressor, ε_3_ or the brain’s trial-by-trial EEG signal reflecting surprise about changes in the statistical structure of the environment. This was the only model-based contrast to achieve FWE-corrected significance, and the only one unconfounded by age. Within the early MMN window (164–195 ms), PSS+ youth showed reduced (more negative) ε_3_ responses relative to PSS– youth over frontal channels. Inspection of the averaged waveforms reveals that this group difference is driven primarily by an exaggerated neural response to high-ε_3_ trials — that is, tones occurring at moments of particularly high environmental surprise, when the model predicts a large update to the brain’s estimate of how rapidly the auditory environment is changing — within the PSS+ group. Responses to low-ε_3_ trials, representing tones that carry little surprise about environmental change, remain comparable between groups (Figure 6). In other words, PSS+ youth might show a selectively amplified neural signature at moments of high environmental unpredictability, rather than a generalised alteration in auditory processing. This might point to a higher expected volatility in less volatile situations in PSS+, in this case, heightened response to sudden unexpected changes in the sounds.

The enhanced ε_3_ negativity in PSS+ youth was prominent during the first stable phase but markedly attenuated during the second (Figure 6.G). This reflects an inflexible inferential strategy rather than a static auditory deficit: the intervening volatile phase triggers an overshoot in posterior uncertainty, leaving an inflated volatility prior that persists into the second stable phase, rendering subsequent deviations less surprising and blunting the ε_3_ update.

This volatility-level signal has a specific pharmacological signature. In a placebo-controlled study of nonclinical volunteers, Weber, Diaconescu et al. (60) showed that the NMDA receptor (NMDAR) antagonist ketamine, which transiently mimics the NMDAR hypofunction implicated in psychosis, selectively altered ε_3_ expression while leaving the low-level sensory prediction error (ε_2_) intact. Because NMDARs mediate the descending predictive signals in the cortical hierarchy, their blockade specifically impairs the updating of higher-level beliefs about how rapidly the environment is changing, without disrupting basic tone-to-tone processing (36). This dissociation identifies ε_3_ as the NMDAR-sensitive component of hierarchical auditory prediction and provides a mechanistic anchor for our finding. The direction, however, differs: ketamine reduced ε_3_ in the MMN window, whereas PSS+ youth showed enhanced ε_3_. This is not necessarily contradictory. As the state-dependent pattern above suggests, an early, exaggerated ε_3_ response and a later, blunted one are two phases of the same process and need not share a single sign; both are consistent with a breakdown in the ability to form a stable internal model of the environment and to capitalise on its regularities (76).

It is important to note, however, that the PSS+ group is diagnostically heterogeneous, carrying a mean of 3.5 comorbid DSM-5 diagnoses, predominantly anxiety and depressive disorders (9). Multiple neuromodulatory mechanisms could plausibly drive the observed ε_3_ pattern: anxiety-driven hypervigilance to environmental change, depression-related alterations in prediction updating, and altered prefrontal-temporal connectivity arising from neurodevelopmental conditions each represent viable contributing pathways that do not require a primary NMDAR-plasticity reduction (46,77,78). Rather than reflecting a single receptor-level deficit, the attenuated frontal ε_3_ signature may represent a common neural endpoint reached via multiple clinical pathways over the course of development (79,80). Determining the specific NMDAR contribution will require pharmacological-challenge designs or molecular imaging (e.g., synaptic vesicle protein 2A PET (81)).

ε_3_ acts as a categorical marker, separating youth who feature any extreme endorsement of psychotic-like experiences on the PRIME revised scale,without tracking continuous symptom severity. This aligns with the Extreme Agreement Index (EAI) identifying a qualitatively distinct profile—characterized by elevated suicidality and functional impairment—rather than merely the high-scoring tail of a distribution (11). In contrast, dimensional analyses revealed an inverse association between unweighted volatility prediction errors (δ_2_) and the General PSS factor at ∼130 ms, alongside a trend-level positive association between posterior uncertainty (ψ_2_) and the Specific_1 factor at 387 ms. Under the HGF framework (37,38), δ_2_ captures reactivity to changing environmental predictability. Because more negative δ_2_ amplitudes in the early MMN window reflect a larger-magnitude response, increasing transdiagnostic burden directly corresponds to an exaggerated neural reaction to environmental change (Figure 7.I–J). Both ε_3_ and δ_2_ thus index a heightened volatility-level response; however, while ε_3_ categorically flags the clinical threshold, δ_2_ scales continuously with general severity. This graded over-reactivity reflects a transdiagnostic hypervigilance to environmental change and, toward the psychosis spectrum, the aberrant assignment of salience to random sensory fluctuations (45,82).

### Volatile-Phase MMN and Social Functioning

The volatile-phase MMN association with GF:Social — replicating a significant finding in non-clinical controls (14) — provides functional grounding for the paradigm’s ecological validity. Prior ERP research links MMN and P300 amplitudes to psychosocial functioning both in healthy adults (83) and across the psychosis spectrum (84–87). Hamilton et al. (85) specifically demonstrated that MMN — but not P300 — was associated with functional disability in schizophrenia, and Thomas et al. (87) modelled a causal pathway from early auditory processing deficits to psychosocial impairment. Our volatile-phase specificity extends this literature in a critical direction by demonstrating that the relevant functional variable is not general MMN amplitude or basic sensory discrimination, but rather the capacity to flexibly adjust predictive coding under changing statistical conditions. This phase-specificity is theoretically coherent: the social world is inherently volatile. Social cognition depends heavily on precision-weighted inference to navigate the shifting intentions of others—a mechanism computationally analogous to the volatility-level inference tracked in our paradigm (88–91). Finally, the directional consistency of this finding across both transdiagnostic youth and healthy adults underscores its robustness.

### Limitations

Several limitations warrant consideration. First, the present analyses are cross-sectional; longitudinal follow-up data from baseline will test whether pwPE–EEG signatures predict change in PSS severity, and whether ε_3_ differentiates youth who subsequently develop a prodromal syndrome from those who remit. Second, the transdiagnostic PSS+ group (mean 3.5 comorbidities, predominantly anxiety and depression) does not permit attribution of the ε_3_ signal to any single diagnostic category or neurobiological substrate. An important extension for future work is thus applying the identical pipeline to established clinical groups and across the psychosis severity continuum (40). Third, because this study utilized a passive listening paradigm, we applied a Bayes-optimal model rather than estimating behavioral parameters for each individual, which may have limited our ability to detect variations between subjects. Finally, despite the large number of subjects in the TAY-CAMH cohort, only a subset of participants received EEG.

## Conclusions

Using mass-univariate ERP analysis and model-based EEG analysis within the HGF framework, this study reveals four principal findings in mental health help-seeking youth. First, age is the dominant modulator of sensory-level prediction error and second, ε_2_ (a measure of sensory-level precision-weighted prediction error) does not distinguish PSS groups. Third and most importantly, volatility-level precision-weighted prediction error (ε_3_) constitutes a categorical, age-independent neural marker of PSS group status. Finally, volatile-context prediction error processing specifically associates with real-world social functioning, replicating findings from non-clinical controls using the identical paradigm used here, complementing the proposed model by Thomas et al. (87) in linking early auditory processing to functional impairments in psychosis. Together, these findings establish that disruption of predictive coding in mental health help-seeking youth is specific to the higher level of the auditory hierarchy where the perception of the environmental volatility is formed. While PSS status is a distinguishing characteristic, it is associated with broad increases in symptom severity, suicidality and functional impairment. Therefore, it likely constitutes a transdiagnostic neural correlate of clinical severity rather than a psychosis-specific risk marker — a distinction with direct implications for computational phenotyping and early intervention strategies.

## Data Availability

All data produced in the present study are available upon reasonable request to the authors.

## Acknowledgements

We extend our thanks to the youth and caregiver participants of the TAY Cohort study. We also thank the clinical staff in Child, Youth and Family Services, the members of the McCain Centre Youth Engagement Initiative, the TAY Cohort Youth Advisory Group, the TAY Cohort Family Advisory Group, and the TAY CAMH Cohort Research Staff for their contributions to this work.

## Author Contributions

**Milad Soltanzadeh** contributed to conceptualization, methodology, data acquisition, formal analysis, software, visualization, and writing – original draft. **Andreea Diaconescu** contributed to conceptualization, methodology, data acquisition, funding acquisition, supervision, and writing – review and editing. **Colleen E. Charlton, John D. Griffiths** and **Zheng Wang** contributed to data curation/formal analysis/software/visualization, and writing – review and editing. **Ore Ogundipe, Denisa Lazar** and **Diya Shah** contributed to data acquisition and data curation, and writing – review and editing. The **TAY Steering Committee** investigators and named youth/family engagement Steering Committee members contributed to study conceptualization, investigation/resources, interpretation of findings, and writing – review and editing.

## Data and Code Availability

Data and analysis code will be made openly available in accordance with FAIR principles upon publication. Pre-processed EEG data and HGF model inversion outputs will be deposited in the Open Science Framework (OSF). Computational modelling code is available via the TAPAS toolbox (*v4.0.0*; https://github.com/ComputationalPsychiatry). Full preprocessing, QC and statistical parametric analysis analysis is available on GitHub: https://github.com/andreeadiaconescu/tay-eeg.git.

