## Supplementary Material for "Volatility-Level Inference Indexes Psychosis Spectrum Symptoms Independent of Age in Transdiagnostic Help-Seeking Youth"

Soltanzadeh[1, 2] et al.

[1] Institute of Medical Science, University of Toronto, Toronto, Ontario, Canada; [2] Krembil Centre for Neuroinformatics, Centre for Addiction and Mental Health, Toronto, Ontario, Canada

### Contents

**S1.** Clinical Details

**S2.** EEG Preprocessing Pipeline

**S3**. Stimulus Timing Verification and Correction

**S4.** Quality Control Criteria and Exclusion Summary

**S5.** HGF Model Specification and Computational Modelling Details

**S6.** Trial Distribution and Data Quality Summary

**S7.** Parametric (GLM) Analysis Pipeline

**S8.** Secondary Results: Full Regressor and Covariate Analyses

**S9.** References

### S1. TAY Clinical Details

#### S1.1 Inclusion and Exclusion Criteria

Inclusion criteria: (i) fluency in English; (ii) accessing health services at CAMH; (iii) age 11–24 years. Exclusion criteria: (i) substance use disorder in the past 6 months; (ii) concomitant major medical or neurological illness; (iii) history of seizure or concussion; (iv) Parent/legal guardian's inability to provide informed consent for the youth.

#### S1.2 Extra Clinical Assessments

Diagnostic evaluations were conducted according to DSM-5 criteria, utilizing the Kiddie Schedule for Affective Disorders and Schizophrenia (K-SADS DSM-5) for participants under 18 years of age, and the Structured Clinical Interview for DSM-5 (SCID-5) for those aged 18 and older (1,2). Suicidality was evaluated using the Columbia-Suicide Severity Rating Scale (C-SSRS) (3). Finally, psychosocial capabilities were quantified using the Global Functioning: Social (GF:Social) and Global Functioning: Role (GF:Role) scales (4).

#### S1.3 PRIME-R Questionnaire Items

Participants filled out the 12-item PRIME-R self-report questionnaire, scored on a 0–6 Likert scale (1). The 12 questions are included below.

***q1.*** *I think that I have felt that there are odd or unusual things going on that I can't explain;* ***q2.*** *I think that I might be able to predict the future;* ***q3.*** *I may have felt that there could possibly be something interrupting or controlling my thoughts, feelings, or actions;* ***q4.*** *I have had the experience of doing something differently because of my superstitions.;* ***q5.*** *I think that I may get confused at times whether something I experience or perceive may be real or may be just part of my imagination or dreams;* ***q6.*** *I* *have thought that it might be possible that other people can read my mind, or that I can read others' minds;* ***q7.*** *I wonder if people may be planning to hurt me or even may be about to hurt me;* ***q8.*** *I believe that I have special natural or supernatural gifts beyond my talents and natural strengths;* ***q9.*** *I think I might feel like my mind is "playing tricks" on me;* ***q10.*** *I have had the experience of hearing faint or clear sounds of people or a person mumbling or talking when there is no one near me;* ***q11.*** *I think that I may hear my own thoughts being said out loud;* ***q12.*** *I have been concerned that I might be "going crazy".*

#### S1.4 Preliminary Baseline Clinical Findings

Baseline data from the Toronto Adolescent and Youth Cohort at the Centre for Addiction and Mental Health (TAY-CAMH) demonstrate that Extreme Agreement Index (EAI)-defined PSS+ youth show are substantially more impaired than PSS– peers across global functioning (Columbia Impairment Scale: d = 0.63; WHODAS: d = 0.74; Global Functioning – Role: d = 0.49), despite a shared tertiary-care context (5). Critically, PSS+ youth show markedly elevated suicidality: roughly twice the rate of past-year suicidal behaviour (26.6% vs. 13.2%) and suicide attempt (15.8% vs. 5.3%), and nearly twice the rate of non-suicidal self-injury (45.1% vs. 26.8%) (5). These differences are not driven by differential substance use, which was statistically indistinguishable between groups and emerged within a context of extensive diagnostic multimorbidity (mean 3.5 DSM-5 diagnoses; predominantly anxiety and depression).

#### S1.5 Bifactor Modeling Metrics

Scree plot analysis indicated a two-factor solution for the specific symptom domains (eigenvalues: 5.74, 1.29, 0.96). The resulting Schmid-Leiman exploratory bifactor model demonstrated excellent overall fit (SRMR = 0.055) and accounted for 58.62% of the total variance. The model identified a robust general factor with substantial loadings across all items ($\lambda$ = 0.48–0.71; see Figure 2.B), which demonstrated excellent total reliability ($\omega_T$ = 0.913).

Dimensionality metrics supported the essential unidimensionality of the scale (Explained Common Variance = 0.698; Percentage of Uncontaminated Correlations = 0.530), with a substantial proportion of variance attributable strictly to the general trait ($\omega_h$ = 0.740). Furthermore, the general factor exhibited excellent construct replicability (H-index = 0.904), confirming a shared, stable transdiagnostic dimension not localized to any single symptom domain (5). Specific_1 factor demonstrated adequate construct replicability (H-index = 0.735) and

### S2. Preprocessing Pipeline

All preprocessing was performed in MATLAB R2023b using SPM12 (*v7771*; http://www.fil.ion.ucl.ac.uk/spm/) and EEGLab (v2025.1.0) (2). The complete preprocessing pipeline is illustrated in Figure S1. Steps were applied identically to all participants, with deviations for individual data quality issues documented below.

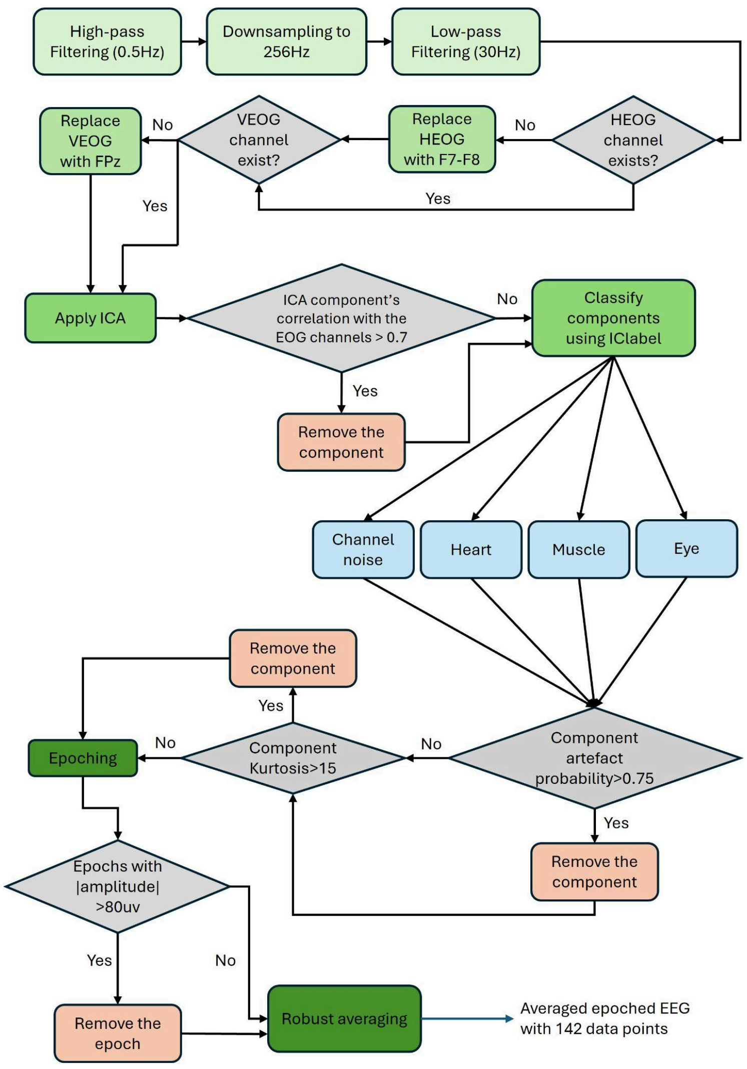


**Figure S1. Preprocessing pipeline flowchart.** Schematic showing the sequential preprocessing steps applied to all participants. Green boxes indicate processing operations; orange diamonds indicate decision points; red boxes indicate exclusion paths. Numbers in parentheses indicate the number of participants affected at each step.

#### S2.1 Referencing, Filtering, and Downsampling

Continuous EEG data were downsampled from 2048 Hz to 256 Hz and re-referenced to the common average of all 64 scalp electrodes. A 5th-order Butterworth band-pass filter was applied with cut-offs at 0.5 Hz (high-pass) and 30 Hz (low-pass). The high-pass cutoff of 0.5 Hz was selected to remove slow drifts while preserving the slow late components of the ERP (e.g., the P300 occurring at 250–350 ms); the low-pass cutoff of 30 Hz removed high-frequency muscle artefacts while retaining all ERP components of interest (3). Filtering was applied to continuous data before epoching to avoid edge effects within epochs.

#### S2.2 Electro-oculogram Preparation

As shown in Figure S2, four Electro-oculogram (EOG) channels were recorded: two vertical (supraorbital and infraorbital to the left eye) and two horizontal (lateral canthi of both eyes). Bipolar VEOG (EX4 - EX8) and HEOG (EX3 - EX7) difference signals were computed from each pair. For participants with flat or missing EOG signals (determined by visual inspection of recording logs and signal screenshots), the following substitutions were applied: VEOG was replaced by channel Fp1 (the nearest frontal electrode most sensitive to vertical eye movements), and HEOG was replaced by the F7 – F8 bipolar difference. Affected participants are listed in Table S1. Independent component analysis (ICA) components that survived the correlation threshold in step S2.3 were additionally screened by ICLabel classification (step S2.4).

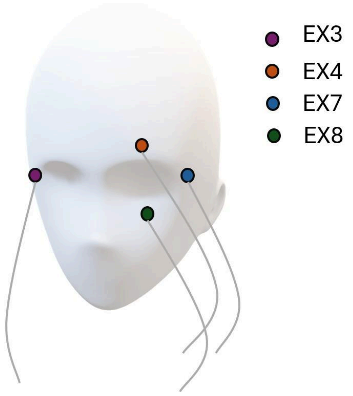


**Figure S2.** EOG Channels Placement. External electrodes EX3, EX4, EX7 and EX8 are placed to record eye blinks and lateral eye movements.

#### S2.3 ICA Decomposition and Artefact Rejection

ICA was performed using the extended Infomax algorithm implemented in EEGLab (2,4). ICA was computed on the full continuous filtered data (before epoching) to maximise the data available for decomposition. Artefact removal proceeded in four sequential steps:

1. **Ocular artefact removal by correlation:** ICA components with Pearson correlation $|r| > 0.70$ with either the VEOG or HEOG channel were removed. This step targets blink and saccade components that are clearly identifiable by their high correlation with recorded eye movement signals.
2. **Automated classification by ICLabel (4)**: Retained components were classified into seven categories (brain, muscle, eye, heart, line noise, channel noise, other) using a pre-trained deep neural network (ICLabel *v1.7*). Components with >75% probability of belonging to any artefact category (muscle, heart, or channel noise) were removed. Eye

components surviving step 1 were additionally removed here if classified as eye artefacts with >75% probability.

3. **Kurtosis-based removal of singular channel noise (5)**: Remaining components with a kurtosis value exceeding 15 were removed. Kurtosis measures the 'peakedness' of the amplitude distribution; values above 15 indicate extreme, non-Gaussian amplitude excursions inconsistent with neural EEG and characteristic of electrode pop or cable artefacts.
4. **Epoch-level amplitude rejection:** After ICA back-projection, epochs were extracted (−100 to 450 ms relative to tone onset) and epochs containing any channel amplitude exceeding ±80 μV relative to the pre-stimulus baseline were rejected.

The quality control threshold for total ICA components removed was set at 26 (the 90th percentile of the distribution across all participants). Participants exceeding this threshold were flagged for manual review; those for whom the retained components still showed unacceptable artefact probability (>25% mean probability of being artefactual across retained components) were excluded from further analysis (see Section S3).

#### S2.4 Epoching and Baseline Correction

Data were epoched into segments spanning −100 to 450 ms relative to each tone onset, yielding one epoch per tone across the 1,800-trial sequence. Baseline correction was applied using the mean amplitude in the −100 to 0 ms pre-stimulus window. The 450 ms post-stimulus epoch length was selected to capture the full P300 component (typically peaking at 250–350 ms) and the reorienting negativity (RON), which can extend to 400 ms, while avoiding overlap with the subsequent tone (inter-stimulus interval = 500 ms).

**Table S1. Number of Participants with Flat EOG Channels**

| Flat channels | Number of participants affected | |
|---|---|---|
| | PSS- | PSS+ |
| VEOG (EX4 & EX8) | 3 | 6 |
| HEOG (EX3 & EX7) | 2 | 3 |

### S3. Stimulus Timing Verification and Correction

Precise alignment of EEG epochs to tone onset times is critical for the mass-univariate parametric analysis, in which trial-by-trial computational regressors must be matched precisely to the EEG epoch from the corresponding trial. A systematic audio delivery latency was identified between the stimulus computer's intended tone onset times and the actual onset times recorded in the EEG trigger channel.

#### S3.1 Source of the Auditory Delivery Latency

The BioSemi ActiveTwo system records EEG and trigger signals in a shared data stream; however, the stimulus presentation software (PsychToolbox-3) generated tone onset triggers via a USB interface, introducing a variable but systematically positive latency relative to the trigger TTL pulse sent to the EEG amplifier. Inspection of the behavioural log files (which recorded intended tone presentation times at millisecond resolution on the stimulus computer) against the EEG trigger channel revealed a stable mean timing offset of 125 ms per trial, consistent across participants and sessions. This offset reflects the audio buffer latency of the USB soundcard and is a known characteristic of this hardware configuration.

##### S3.2 Correction Procedure

Stimulus onset times were verified and aligned on a trial-by-trial basis using event timestamps recorded in the parallel behavioural log file, a standard calibration procedure for BioSemi ActiveTwo recordings. For each participant, the per-trial offset between the behavioural log timestamp and the EEG trigger was computed (see *tayeeg_delay_adjustment.m* in the GitHub repository). EEG epochs were then extracted relative to the corrected trigger times. The correction was validated by comparing the peak latency of the grand-average MMN before and after correction: the corrected MMN peaked at ~180 ms, consistent with canonical MMN latency in the literature (6–8).

#### S4. Quality Control Criteria and Exclusion Summary

Quality control (QC) was applied after preprocessing to identify participants whose EEG data did not meet minimum standards for reliable mass-univariate analysis. Five pre-specified QC criteria were evaluated, each with a rationale:

##### S4.1 QC Criteria

1. Number of artefact-free trials < 1,000. **Rationale:** The mass-univariate parametric analysis regresses trial-wise computational quantities against EEG amplitude, requiring sufficient trial-by-trial variance. Simulations by Charlton et al. (8) and Weber, Diaconescu et al. (7) indicate that >1,000 artefact-free trials are required for stable HGF regressor estimation. The median number of artefact-free trials across retained participants was well above this threshold (see Table S3).

2. Number of bad channels > 5. **Rationale:** Bad channels were interpolated prior to analysis. Interpolating more than 5 of 64 channels (>8%) introduces spatial smoothing that could obscure the topographic specificity of group differences.
3. Number of removed ICA components > 26. **Rationale:** This threshold corresponds to the 90th percentile of the distribution across all participants (see Table S3). Participants requiring removal of >26 components had residual artefact levels after ICA that risked contaminating the single-trial parametric analysis.
4. Mean probability of retained components being artefactual > 25%. **Rationale:** Even after ICA component removal, retained components may carry residual artefact probability. A mean artefact probability >25% across retained components indicates that the ICA decomposition was insufficiently clean for single-trial analysis.
5. Behavioral hit rate on visual distraction task < 50%. **Rationale:** The visual task was designed to direct attention away from the tones, ensuring passive auditory processing. A hit rate below 50% (chance level for a 2-alternative task) may indicate that the participant was attending to or ignoring the tones rather than performing the visual task, potentially altering the auditory ERP profile.

#### S4.2 Exclusion Summary

Of 149 participants who completed the EEG session and met initial inclusion criteria, 18 were excluded based on QC, yielding a final analytic sample of N=131. Figure S3 provides a per-criterion breakdown of exclusions.

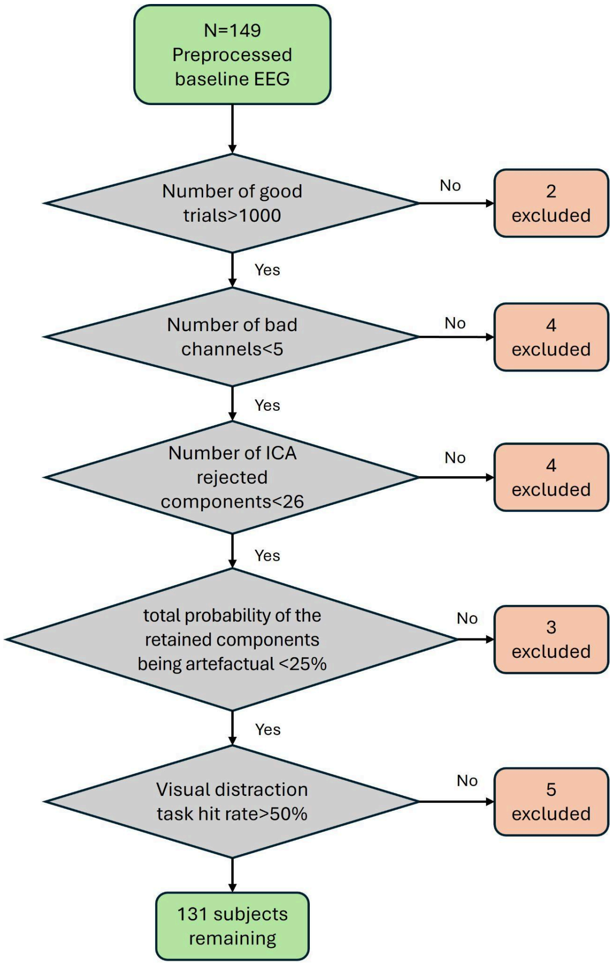


**Figure S3. Quality control flowchart and the exclusion summary.**

### S5. HGF Model Specification and Computational Modelling Details

Trial-by-trial computational quantities were derived using the Hierarchical Gaussian Filter (HGF, *version 6* (9)) implemented via the *TAPAS* toolbox (*v4.0.0*; https://github.com/ComputationalPsychiatry) (10). The following subsections describe the model structure, parameter settings, and regressor extraction procedure in full.

#### S5.1 Model Structure

The HGF is a generic hierarchical Bayesian model of learning under uncertainty (11,12). In the present auditory oddball context, we employed the binary three-level HGF (tapas_ehgf_binary), which models an observer's implicit learning of a binary tone sequence. The model assumes the observer infers three hierarchically coupled hidden states on each trial k:

- **Level 1 ($x_1$):** The observed binary tone identity (high tone = 1, low tone = 0).
- **Level 2 ($x_2$):** The current tone tendency — the observer's belief about the probability of hearing the high tone. This state evolves as a Gaussian random walk with step-size controlled by level 3.
- **Level 3 ($x_3$):** The log-volatility of the environment — the observer's belief about how rapidly the tone tendency is changing. This top-level state evolves as a slower Gaussian random walk, tracking the overall pace of environmental change.

Beliefs at each level are characterised by their posterior mean ($\mu_i$) and variance ($\sigma_i$). After each tone, beliefs are updated via precision-weighted prediction errors according to:

$$\Delta\mu_i^{(k)} \propto (\hat{\pi}_{i-1}^{(k)} / \pi_i^{(k)}) \times \delta_{i-1}^{(k)}$$

where $\delta_{i-1}^{(k)}$ is the unsigned prediction error from the level below, $\hat{\pi}_{i-1}^{(k)}$ is the precision of the prediction about the level below (sensory precision), and $\pi_i^{(k)}$ is the precision of the current belief at level i. The ratio of precisions functions as a dynamic learning rate, scaling the influence of prediction errors on belief updating.

#### S5.2 Parameter Settings: Bayes-Optimal Observer

Since the auditory oddball paradigm does not require behavioural responses, HGF parameters could not be estimated from participant behaviour. Consistent with prior applications of the HGF to MMN paradigms (7,8,13), participants were modelled as surprise-minimising Bayesian observers. For each participant's specific tone sequence, perceptual parameters were optimised to minimise the total surprise (negative log model evidence) evoked by the experienced tone sequence, using the tapas_fitModel function with the tapas_bayes_optimal_whatworld pseudoresponse model. This procedure yields slightly different parameter values for each participant's session, reflecting their specific trial sequence. Prior and posterior values of the HGF parameters are reported in Table S2.

**Table S2.**

| Parameter | Description | Prior | Posterior |
|---|---|---|---|
| **$\omega_2$ (tonic volatility, level 2)** | Baseline learning rate independent of online volatility estimation | -3 | -1.91 |
| **$\omega_3$ (tonic volatility, level 3)** | Speed of change in volatility estimates | 2 | 1.95 |
| **$\mu_3^{(0)}$ (initial volatility belief)** | Starting value for log-volatility estimate | 1 | N/A |
| **$\sigma_3^{(0)}$ (initial volatility uncertainty)** | Starting uncertainty about log-volatility | 0 | N/A |

#### S5.3 Computational Regressors

Six trial-by-trial computational quantities were extracted from the HGF inversion for use as EEG regressors:

- $\varepsilon_2$ (epsilon2): Low-level sensory precision-weighted prediction error. Reflects the influence of sensory surprise about the tone identity on updating the level-2 belief about tone tendency. $\varepsilon_2 = (\hat{\pi}_1^{(k)} / \pi_2^{(k)}) \times \delta_1^{(k)}$.
- $\varepsilon_3$ (epsilon3): Volatility-level precision-weighted prediction error. Reflects the influence of precision-weighted surprise about the tone tendency on updating the level-3 volatility estimate. $\varepsilon_3 = (\hat{\pi}_2^{(k)} / \pi_3^{(k)}) \times \delta_2^{(k)}$.
- $\delta_1$ (delta1): Unsigned low-level prediction error. The raw, unweighted discrepancy between the predicted and actual tone identity at level 1, before precision-weighting.
- $\delta_2$ (delta2): Unsigned high-level prediction error. The raw, unweighted discrepancy between the predicted and actual tone tendency at level 2, before precision-weighting. Reflects the magnitude of belief-updating at the volatility level independently of precision.
- $\psi_2$ (psi2): Posterior uncertainty at level 2. The posterior variance $\sigma_2^{(k)}$ of the belief about tone tendency, reflecting accumulated uncertainty about the current strength of tone-to-tone associations.
- $\psi_3$ (psi3): Posterior uncertainty at level 3. The posterior variance $\sigma_3^{(k)}$ of the belief about log-volatility, reflecting uncertainty about the current rate of environmental change.

##### S5.4 Regressor Collinearity

Mean pairwise Pearson correlations between regressors (computed across all trials, averaged across participants) are shown in Figure S4. The collinearity structure replicates that reported by Charlton et al. (8) ($r=0.087\pm0.008$ for $\delta_1/\psi_2$; $r=0.118\pm0.006$ for $\delta_2/\psi_3$ in their sample), confirming that the HGF implementation yields comparable regressor properties in this clinical youth sample.

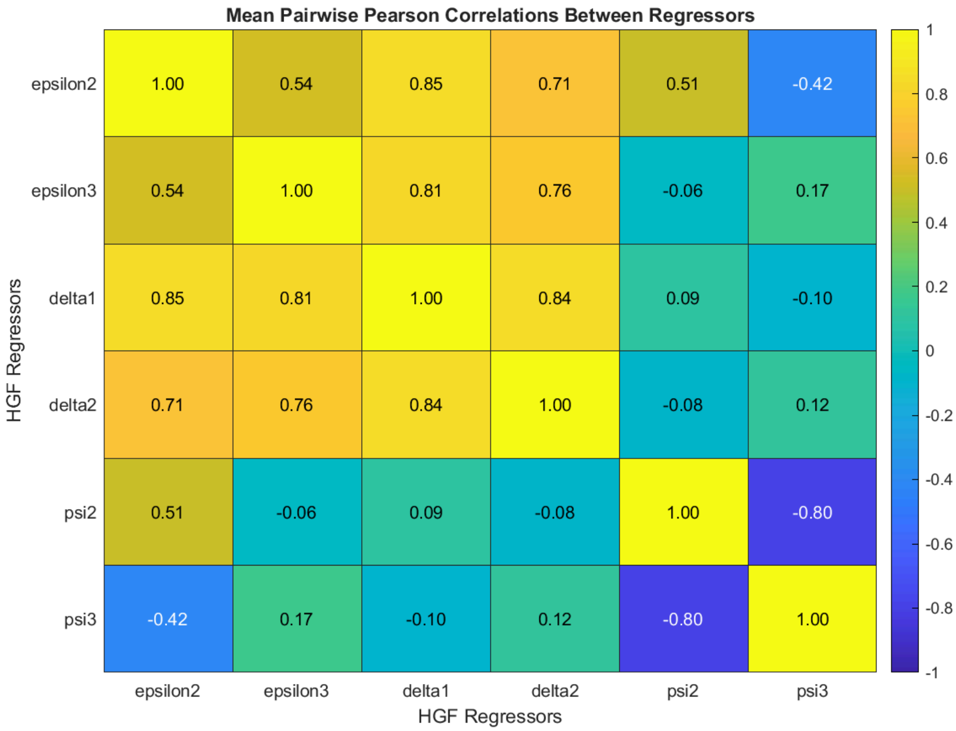


**Figure S4. Mean pairwise Pearson's correlation between HGF regressors.**

### S6. Trial Distribution and Data Quality Summary

To verify that data quality did not bias our group-level EEG contrasts, we assessed trial counts and quality metrics for the PSS+ and PSS– groups (see Table S3). Despite a significant between-group difference in the raw number of artifact-free trials, the final count of trials used for averaging was consistent across both groups. This ensures that the signal-to-noise ratio was balanced and the final results were not confounded by data retention disparities.

##### Table S3. Trial Distribution and Data Quality Summary Across groups

| Metric | | | PSS– (N=66; mean ± SD) | PSS+ (N=65; mean ± SD) | Statistic |
|---|---|---|---|---|---|
| **Number of initial trials** | | | 1799±2.91 | 1796±25.57 | p-value=.34 |
| **Number of rejected epochs*** | | | 101.70±137.71 | 51.49±73.46 | p-value=.01 |
| **Number of bad channels** | | | 0.04±0.21 | 0.38±2.66 | p-value=.08 |
| **Number of artefact-free trials*** | | | 1697±137.92 | 1744±75.46 | p-value=.01 |
| **Number of final averaged trials**[1] | **Standard** | **Oddball** | 105.95±0.21 | 105.8±1.49 | p-value=.41 |
| | | **Stable** | 50.95±0.21 | 50.98±0.12 | p-value=.32 |
| | | **Volatile** | 55±0.00 | 54.81±1.49 | p-value=.31 |
| | **Deviant** | **Oddball** | 119±0.00 | 118.80±1.61 | p-value=.31 |
| | | **Stable** | 55±0.00 | 55±0.00 | same |
| | | **Volatile** | 64±0.00 | 63.8±1.61 | p-value=.31 |
| **ICA** | **Number of components removed** | | 13.82±4.70 | 12.68±4.19 | p-value=.14 |
| | **Mean artefact probability of retained components (%)** | | 0.10±0.05 | 0.10±0.05 | p-value=.72 |
| **Visual task** | **Visual task hit rate (%)** | | 87.28±21.22 | 89.65±13.65 | p-value=.43 |
| | **Visual task reaction time (s)** | | 0.76±0.12 | 0.75±0.14 | p-value=.79 |

* Asterisks indicate significant differences ($p_{value} < 0.05$)

[1] The standard/deviant trials are predefined by design for all the subjects. The minor differences between the two groups come from the rejected artefactual trials.

### S7. GLM Analysis Pipeline

Single-trial EEG data were analysed using a parametric general linear model (GLM) approach implemented in SPM12, following the pipeline established by Weber, Diaconescu et al. (7) and subsequently applied in Charlton et al. (8) and Hauke et al. (13).

#### S7.1 Scalp Image Construction

For each participant, preprocessed EEG difference waveforms were converted into 2D scalp images (time × sensor space). Each epoch was mapped to a 2D sensor grid using linear interpolation across the 64 electrode positions, yielding images with a voxel size of 4.3 mm × 5.4 mm × 2.0 ms (spatial × spatial × temporal dimensions). Images were smoothed with an isotropic Gaussian kernel (full-width at half-maximum: FWHM = 16 × 16 mm spatially) to satisfy the assumptions of Gaussian random field theory used for multiple comparison correction (14). All images were constructed across the peristimulus time window of interest (0–450 ms), but statistical analyses were restricted to 100–400 ms to capture MMN, P300, and RON components while reducing the number of comparisons.

#### S7.2 First-Level (Subject-Level) GLM

For each participant and each session, a general linear model was specified with the following regressors:

- An intercept term (modelling the mean EEG amplitude across all trials).
- Z-standardised computational trajectory regressors: $\varepsilon_2$, $\varepsilon_3$, $\delta_1$, $\delta_2$, $\psi_2$, $\psi_3$ (z-standardised across the trial sequence for each participant separately to ensure

comparability across participants and sessions). Entries corresponding to rejected trials were removed from the regressor vectors before GLM estimation.

#### S7.3 Second-Level (Group-Level) GLM

First-level images served as input to the second-level group analyses. For each computational quantity, a separate second-level GLM was specified. The following second-level models were estimated:

1. One-sample F-test across all participants: tests the null hypothesis that the difference waveform is zero across the sample (main effect of each regressor).
2. Two-sample t-tests (PSS+ vs. PSS–): tests group differences in the β images for each regressor. Age (z-scored) and sex (dummy-coded) were entered as nuisance covariates of no interest.
3. Simple regression with PRIME-R total score and bifactor factor scores (General, Specific1, Specific2) as continuous covariates.
4. Simple regression with GF:Social and GF:Role scores as continuous covariates, estimated separately for the full paradigm, stable-phase, and volatile-phase difference waveforms.

#### S7.4 Statistical Inference

For all analyses, significant effects were inferred using thresholded F- or T-statistical parametric maps (SPMs) with a cluster-defining threshold of $p<0.001$ (uncorrected) and family-wise error (FWE) correction applied at the cluster level ($p<0.05$) using Gaussian random field theory (15). This two-step inference controls the family-wise error rate across the joint time × sensor search volume while maintaining sensitivity to spatiotemporally extended effects.. Results are reported as peak F- or T-values, Z-equivalents ($Z_E$) and FWE-corrected p-values at both peak and cluster levels, together with peak voxel coordinates in time (ms) and sensor space (mm). Six contrast families were evaluated:

(1) Main effect of oddball MMN (deviant>standard).

(2) Stable-phase vs. volatile-phase MMN (paired contrast).

(3) Age as a covariate across all regressors (oddball ERP, $\varepsilon_2$, $\varepsilon_3$, $\delta_1$, $\delta_2$, $\psi_2$, $\psi_3$).

(4) PSS+>PSS– and PSS+<PSS– for: oddball ERP, stable-phase, volatile-phase, $\varepsilon_2$, $\varepsilon_3$, $\delta_1$, $\delta_2$, $\psi_2$, $\psi_3$.

(5) Continuous PRIME-R total score and bifactor scores (General, Unusual thought content, and Grandiosity/superstitions) as parametric covariates.

(6) GF:Social and GF:Role as continuous covariates, separately for the full paradigm, stable-phase, and volatile-phase MMN, following Charlton et al. (6).

#### S7.5 Expression of pwPE and precision regressors

All six computational regressors showed significant main effects in the full sample: $\varepsilon_2$ peaked at ~180ms frontocentral and occipital; $\varepsilon_3$ peaked at ~180 ms frontocentral and occipital; $\delta_1$ and $\delta_2$ peaked at ~180 and ~300 ms frontocentral; ψ2 peaked at ~200, 300 and 340 ms central; ψ3 peaked at ~200 and 300 ms central. This timing and topographic structure overlaps with the regressor expression reported by Charlton et al. (6) in healthy controls and Weber, Diaconescu et al. (7) in their ketamine study, validating the HGF implementation in this clinical youth sample. See Table 3 in the main text for regressors' main effect statistics.

### S8. Secondary Results: Full Regressor and Covariate Analyses

This section reports the complete statistical results for all regressors and contrasts not presented in the main text, providing full transparency for replication and meta-analysis. All statistics are from the post-QC sample (N=131; PSS–=66, PSS+=65) with age and sex as nuisance covariates in all group comparisons.

#### S8.1 $\varepsilon_2$ Group Contrasts and Age Confound

The $\varepsilon_2$ regressor did not yield FWE-corrected group differences between PSS+ and PSS– youth after controlling for age and sex:

- Group contrasts
  - PSS+ < PSS–: $T(126)=3.82$; $p_{uncorr}<0.001$; $p_{FWE\text{-}peak}=0.11$; $k_E=122$; $p_{FWE\text{-}clus}=0.20$; peak: (51, 40, 262 ms),
  - PSS+ > PSS–: No suprathreshold clusters.
- Age effect on $\varepsilon_2$: $F(1,126)=16.03$; $p_{uncorr}<0.001$; $p_{FWE\text{-}peak}=0.13$; $k_E=122$; $p_{FWE\text{-}clus}=0.099$; peak: (38, -62, 223 ms)

#### S8.2 $\delta_1$ Results

- Group contrast (not significant)
  - PSS+ < PSS–: $T(126)=3.22$; $p_{uncorr}= 0.001$; $p_{FWE\text{-}peak}= 0.396$; $k_E=13$; $p_{FWE\text{-}clus}= 0.422$; peak: (−34, 2, 395 ms),
  - PSS+ > PSS–: No suprathreshold clusters.

- Age effect on $\delta_1$: Reported in main text (see Figure 5)

#### S8.3 $\delta_2$ Results

- Group contrast (not significant)
    - PSS+ < PSS–: $T(126)=3.71$; $p_{uncorr}< 0.001$; $p_{FWE\text{-}peak}= 0.160$; $k_E=276$; $p_{FWE\text{-}clus}= 0.089$; peak: (60, 34, 309 ms)
    - PSS+ > PSS–: $T(126)=3.21$; $p_{uncorr}= 0.001$; $p_{FWE\text{-}peak}= 0.483$; $k_E=7$; $p_{FWE\text{-}clus}= 0.463$; peak: (−34, -57, 156 ms)
- Age effect on $\delta_2$: Reported in main text (see Figure 5)

#### S8.4 $\psi_2$ Results

- Group contrasts (not significant)
    - PSS+ < PSS–: $T(126)=3.16$; $p_{uncorr}=0.001$; $p_{FWE\text{-}peak}=0.516$; $k_E=1$; $p_{FWE\text{-}clus}=0.501$; peak: (-60, -68, 316ms),
    - PSS+ > PSS–: No suprathreshold clusters.
- Age effect on $\psi_2$: $F(1,126)=15.91$; $p_{uncorr}<0.001$; $p_{FWE\text{-}peak}=0.14$; $k_E=280$; $p_{FWE\text{-}clus}=0.081$; peak: (42, -62, 137 ms)

#### S8.5 $\psi_3$ Results

- Group contrasts (not significant)
    - PSS+ < PSS–: $T(126)=3.43$; $p_{uncorr}<0.001$; $p_{FWE\text{-}peak}=0.312$; $k_E=20$; $p_{FWE\text{-}clus}=0.406$; peak: (-60, -68, 328 ms),
    - PSS+ > PSS–: No suprathreshold clusters.

- Age effect on $\psi_3$: $F(1,126)=12.68$; $p_{uncorr}=0.001$; $p_{FWE\text{-}peak}=0.41$; $k_E=15$; $p_{FWE\text{-}clus}=0.467$; peak: (-34, 34, 355 ms)

#### S8.6 Full $\varepsilon_3$ Group Contrast Statistics

For transparency, the complete $\varepsilon_3$ group contrast statistics are reported here alongside the main text values. The main text reports significant effects for PSS+ < PSS– [$T(126)=4.30$; $p_{FWE\text{-}peak}=0.024$; $k_E=379$; $p_{FWE\text{-}clus}=0.063$; peak: (−21,−3, 180 ms)] after controlling for age and sex. Earlier analyses prior to covariate inclusion yielded T=4.49 (PSS+ < PSS–, $p_{FWE\text{-}peak}=0.016$, $p_{FWE\text{-}clus}=0.028$) and T=3.92 (PSS+ > PSS–, $p_{FWE\text{-}peak}=0.095$; $p_{FWE\text{-}clus}=0.045$). The post-covariate statistics are the appropriate values and are reported consistently in the main text and Table 2.

#### S8.7 Bifactor Factor Score Covariate Results

Results for all bifactor factor score covariate analyses not included in the main text are provided here for completeness:

- General factor
  - $\delta_2$: Significant inverse association [$T=4.02$; $p_{FWE\text{-}peak}=.065$; $k_E=471$; $p_{FWE\text{-}clus}=0.04$; peak: (-8, 8, 137)]. See Figure 7 in the main text..
  - Other regressors: No significant association.
- Specific1
  - Trend-level associations for $\psi_2$ at ~380 ms parietal reported in main text
  - $\psi_3$ at ~250 ms temporal [$T(126)=3.90$; $p_{uncorr}<0.001$; $p_{FWE\text{-}peak}=0.098$; $k_E=186$; $p_{FWE\text{-}clus}=0.141$; peak: (55, -25, 254 ms)];
  - All regressors: no significant association
